# Prenatal exposure to semi-volatile organic compounds and early childhood sleep duration

**DOI:** 10.64898/2026.11.25.26368881

**Authors:** Natalie R. Binczewski, Jillian H. Hurst, Jessica R. Lunsford-Avery, Rebecca M. Hoehn, Elizabeth F. Boxer, Heather M. Stapleton, Kate Hoffman

## Abstract

**Background:** Adequate sleep duration is critical for early childhood development; however, environmental determinants of sleep during this period remain understudied.

**Objective:** We evaluated associations between prenatal exposure to semi-volatile organic compounds (SVOCs) and parent-reported sleep duration among 147 children 2-24 months of age.

**Methods:** Mothers in Project HOPE 1000, a prospective pregnancy cohort in central North Carolina, wore silicone wristbands for 5 days during each trimester, and 117 SVOCs were quantified. Generalized linear mixed-effects models were adjusted for child age, race, ethnicity, and an exposure-age interaction term.

**Results:** At least 70% of wristbands had detectable levels of 36 SVOCs. Among 2-month-olds, higher prenatal exposure to polycyclic aromatic hydrocarbons, phthalates, and organophosphate esters was associated with decreased sleep duration across daytime, nighttime, and total sleep over a 24-hour period (e.g., benzyl butyl phthalate exposure was associated with lower expected nighttime sleep duration (Multiplicative Change: 0.77, 95% Confidence Interval: 0.61, 0.95). Results among 2-month-olds were similar when exposure was assessed as a mixture. Associations were largely null at older ages across all sleep outcomes.

**Significance:** Prenatal SVOC exposure may have significant impacts on sleep duration during early infancy. Future studies should assess how these differences relate to broader measures of sleep health.

**Impact:** Few studies have investigated impacts of prenatal SVOC exposure on sleep health outcomes in infancy and toddlerhood. We found that prenatal exposure to SVOCs was associated with lower expected sleep duration among 2-month-olds, providing some evidence of prenatal exposure effects persisting into the postnatal period. These associations were also observed when SVOC exposure was considered as a mixture, highlighting the potential for these commonly co-occurring environmental chemicals to jointly adversely impact sleep duration in early infancy.

## Introduction

Achieving sufficient sleep during childhood is crucial for supporting healthy development and may predict later-life health; however, an estimated 44% of children are not consistently achieving the recommended amount of sleep for their age.^1–5^ Sleep is regulated by homeostatic and circadian processes, which are initially established in utero. During pregnancy, the fetal circadian system receives external time cues via maternal hormones crossing the placenta.^6,7^ Cyclical changes in melatonin and cortisol secretion are important for circadian rhythm regulation and entrainment.^4^ Because these systems are established in utero and continue to mature throughout early childhood, there is a need to identify how external signals may impact healthy developmental trajectories.^6^

Environmental risk factors have been associated with sleep problems in adults, yet evidence for environmental influences on childhood sleep remains sparse. Limited research has investigated impacts of urbanicity, second hand smoke, noise, and artificial light at night on childhood sleep health outcomes (e.g., sleep disorders, sleep duration and quality).^8–12^ More recent data suggests associations between exposure to semi-volatile organic compounds (SVOCs) and increased odds of sleep problems, disturbances, and impairment in children between 6 months and 8 years of age.^13–17^ A biologically plausible mechanism for these associations involves disruption of circadian regulation. Because many SVOCs have endocrine-disrupting properties, prenatal exposure may alter hormonal pathways involved in circadian regulation. Circadian dysfunction, in turn, may significantly impact key aspects of sleep health— such as sleep duration—by disrupting the timing and consolidation of sleep.^6^

Elucidating associations between SVOCs and sleep may be particularly critical in infancy and toddlerhood, a developmental period characterized by rapid development in circadian and homeostatic sleep processes.^18^ In this study, we evaluated associations between prenatal SVOC exposure and repeated measures of sleep duration (daytime, nighttime, and total sleep in a 24-hour period) in children between 2- and 24-months of age in a North Carolina pregnancy cohort. We hypothesized that higher exposure to SVOCs is associated with longer daytime sleep and shorter nighttime sleep, representative of a lack of consolidated sleep patterns, and that these associations differ across age groups.

## Methods

### Study Population

Mother-child pairs included in this study were participants enrolled in Project HOPE 1000, a prospective birth cohort based in central North Carolina. Briefly, mothers were recruited during early pregnancy visits between 2019 and 2024, with children followed until 5 years of age. Variables related to pregnancy factors and sociodemographic characteristics were abstracted from electronic health record (EHR) data and surveys administered at study visits. This study was approved by the Duke University Health System Institutional Review Board (Pro00100000; Pro00113307).

### Wristband Analysis for SVOC Exposures

Mothers wore silicone wristbands for 5 days in at least one trimester of pregnancy. Wristband samples were collected and exposure to 117 SVOCs was quantified using gas chromatography high resolution mass spectrometry (GC-HRMS). Chemical names and abbreviations are available in **Supplemental Table 1**. SVOCs were quantified using a single measurement, with the exception of tris (1-chloro-isopropyl) phosphate (TCPP) reported as the sum of its three major isomers. Wristband preparation, collection, and analysis details are available in Hoehn et al. (2027). Briefly, portions of each wristband were cut (~0.5 g), spiked with isotopically labeled internal standards, and solvent extracted with hexane:ethyl acetate. Extracts were further purified with Florisil, filtered, and analyzed using Q exactive and Exactive GC-HRMS systems (Thermo Scientific) operated in electron ionization (EI) negative chemical ionization (NCI) modes, respectively. Field blanks (non-worn wristbands) and laboratory processing blanks (solvent only) were included in all laboratory batches for quality assurance and quality control. Measurements were blank corrected and values below the method detection limit (MDL) were imputed using mass-adjusted MDLs divided by 2.

### Sleep Duration Outcomes

Child sleep duration data were collected via parent-report surveys administered at study visits. Assessments were conducted at 12-month and 24-month visits; children were also randomized to complete assessments at one additional visit at 2-months, 4-months, or 6-months. Mothers indicated the average amount of daytime and nighttime sleep achieved by their child by selecting from predefined sleep duration intervals. Children were assigned to the midpoint of the numeric interval to represent daytime and nighttime sleep durations. To calculate total sleep across a 24-hour period, daytime and nighttime sleep durations were summed.

### Statistical Analysis

Descriptive statistics were calculated for all 117 SVOCs quantified via GC-HRMS to assess data distribution. SVOCs detected in at least 70% of wristband samplers were included in further statistical analyses. Concentrations of permethrin quantified as individual isomers were summed prior to further analysis due to high correlations and inherent co-occurrence in commercial products. Concentrations of SVOCs followed a non-normal distribution (Shapiro-Wilk test), necessitating the use of nonparametric statistics and log10 transformations in subsequent modeling and analysis. Intraclass correlation coefficients (ICC) were calculated to assess the consistency of SVOC exposures for each mother across pregnancy. Individual chemical exposures were largely consistent over time (ICC > 0.5 for 32/36 SVOCs), and therefore prenatal exposure to SVOCs was ultimately defined as the average concentration in all available trimester wristband samplers (1-3 samplers per participant; **Supplemental Table 2**). Spearman correlations between average exposures to individual SVOCs were calculated to evaluate co-exposure patterns.

Generalized linear mixed effects models (Poisson distribution with a log link) with a random intercept for each participant were used to assess log10-transformed average SVOC exposures as predictors of sleep duration (nighttime, daytime, and total sleep across a 24-hour period). To prevent overfitting given the limited sample size, models were adjusted for age at assessment (discrete variable) and race and ethnicity (Non-Hispanic White and All Other Race and Ethnicity Groups), due to associations with both early childhood sleep duration and SVOC exposure.^19,20^ Given the exploratory nature of this analysis and a priori hypothesis that associations between exposure and sleep duration vary across developmental stages, models were fit with an exposure-age interaction term to assess effect modification. Interaction terms were retained in all models regardless of statistical significance. We present results as the multiplicative change in expected sleep duration per 10-fold increase in exposure and 95% confidence intervals.

SVOC exposures were further assessed as a mixture using an average z-score index. The log10 average concentrations were standardized to z-score distributions specific to each individual SVOC. Z-scores were averaged to represent prenatal SVOC exposure burden relative to the study population across the 36 frequently detected SVOCs. Generalized linear mixed effects models were used as described above, with the exposure variable defined as the average z-score. Results are presented for a one standard deviation increase in average SVOC exposure index. All analyses were performed in R (version 4.5.1).^21^

## Results

### Study population and SVOC detection during pregnancy

147 mother-infant pairs had silicone wristbands available from at least one trimester of pregnancy and sleep duration information for at least one time point. Of these, 45 contributed two and 12 contributed three repeated measurements of sleep duration (**Table 1**). Mothers were more frequently Non-Hispanic White with a high household income, post-graduate education, and private insurance. Of the 117 SVOCs assessed in first (n=49), second (n=138), and third (n=136) trimester wristbands, 36 compounds were detected in at least 70% of samples across pregnancy (**Supplemental Table 3**). Average exposures across pregnancy were generally highest among phthalates and organophosphate esters (OPE) chemical classes (**Supplemental Table 4**). Correlations were generally moderately positive between chemicals, with the strongest correlations observed within the same chemical class (e.g., among polycyclic aromatic hydrocarbons (PAHs), phthalates, and brominated flame retardants (BFRs)) (**Supplemental Table 5)**.

**Table 1.**
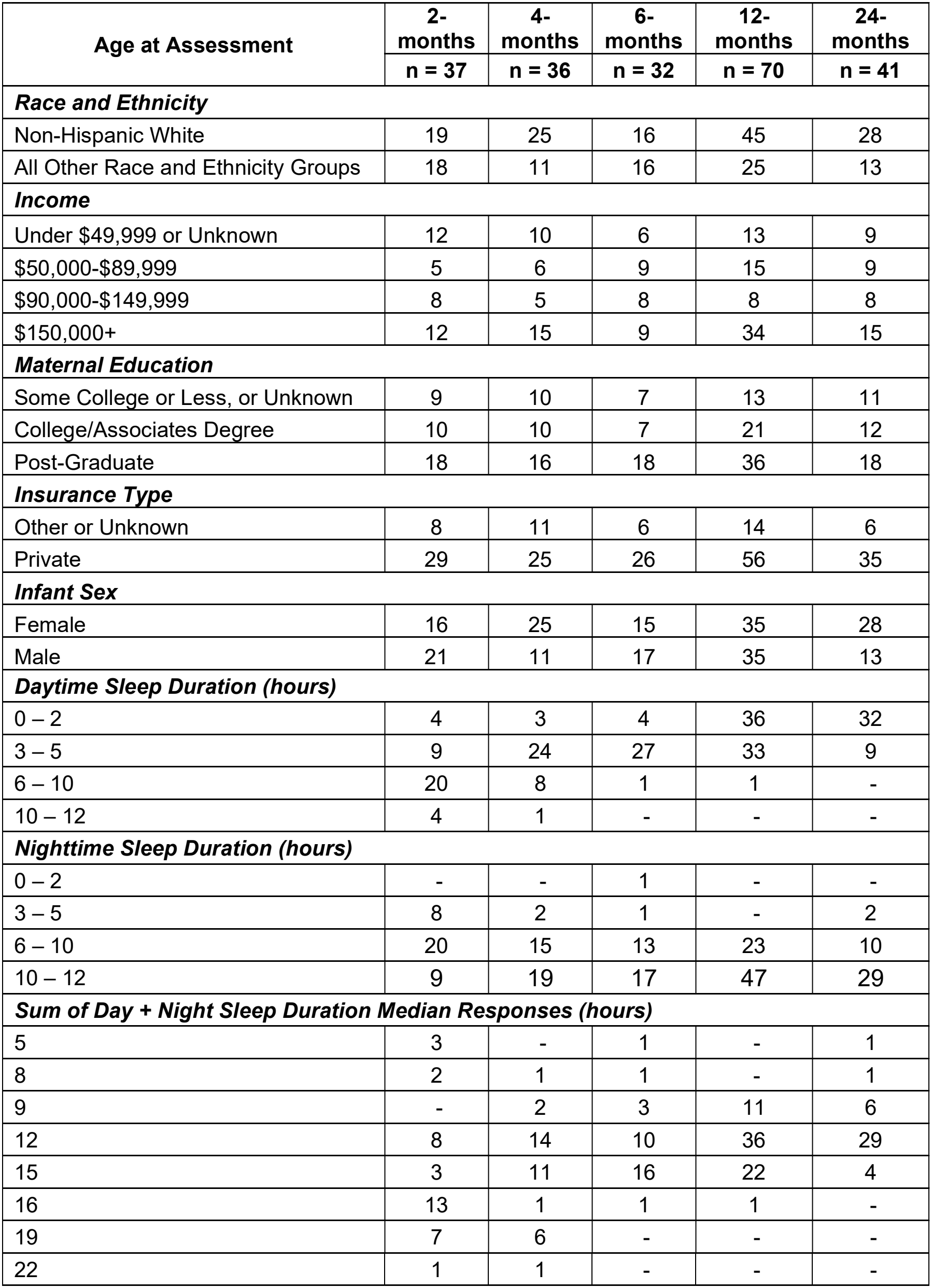
Study population characteristics and distribution of sleep duration outcomes.

### Daytime sleep duration

Among 2-month-olds, exposure to SVOCs across chemical classes (PAHs, phthalates, and OPEs) was associated with decreased expected daytime sleep duration (**Figure 1; Supplemental Table 6)**. Significant associations with daytime sleep duration ranged from 27% to 62% lower expected sleep per 10-fold increase in exposure to benzyl butyl phthalate (BBP) and for nonylphenol (NP), respectively. In 4-month olds, di-ethyl phthalate (DEP) was associated with a 43% (95% CI: 0.33, 0.97) decrease in expected daytime sleep duration. Associations with other SVOCs and daytime sleep duration at 4-months and older were largely positive across chemical classes, albeit non-significant. We also evaluated for associations between total SVOC exposure, evaluated as an average z-score measure, and daytime sleep duration (**Supplemental Table 7**). A one-unit increase in total SVOC exposure was associated with a 23% reduction (95% CI: 0.59, 1.00) in expected daytime sleep duration in 2-month-olds. We did not identify any associations between total SVOC exposure and daytime sleep duration at other ages.

**Figure 1.**
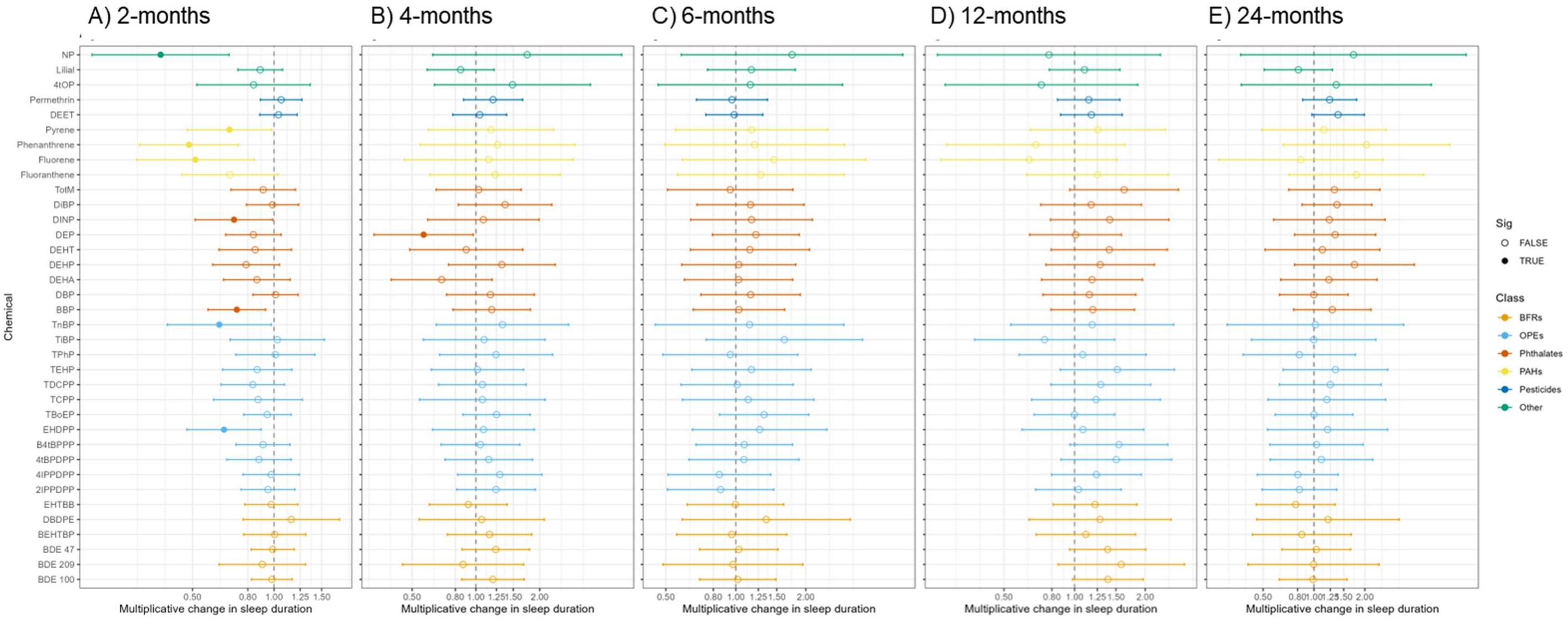
Age-specific estimates from generalized linear mixed effects models with an exposure-age interaction term examining associations between SVOC exposure and daytime sleep at A) 2-months, B) 4-months, C) 6-months, D) 12-months, and E) 24-months old.

### Nighttime sleep duration

Higher exposure to some phthalates, fluorene, and lilial was associated with significantly decreased expected nighttime sleep duration in 2-month-olds (**Figure 2; Supplemental Table 6**). Associations with decabromodiphenyl ethane (DBDPE) and 2-isopropylphenyl diphenyl phosphate (2IPPDPP) were positively associated with nighttime sleep. We did not identify any significant associations between prenatal SVOC exposure and nighttime sleep duration at other ages. Total SVOC exposure was not significantly associated with nighttime sleep duration; however, there was a negative though non-significant association between total SVOCs and decreased nighttime sleep duration among 2-month-olds (e^β^=0.81, 95% CI: 0.64, 1.02) (**Supplemental Table 7**).

**Figure 2.**
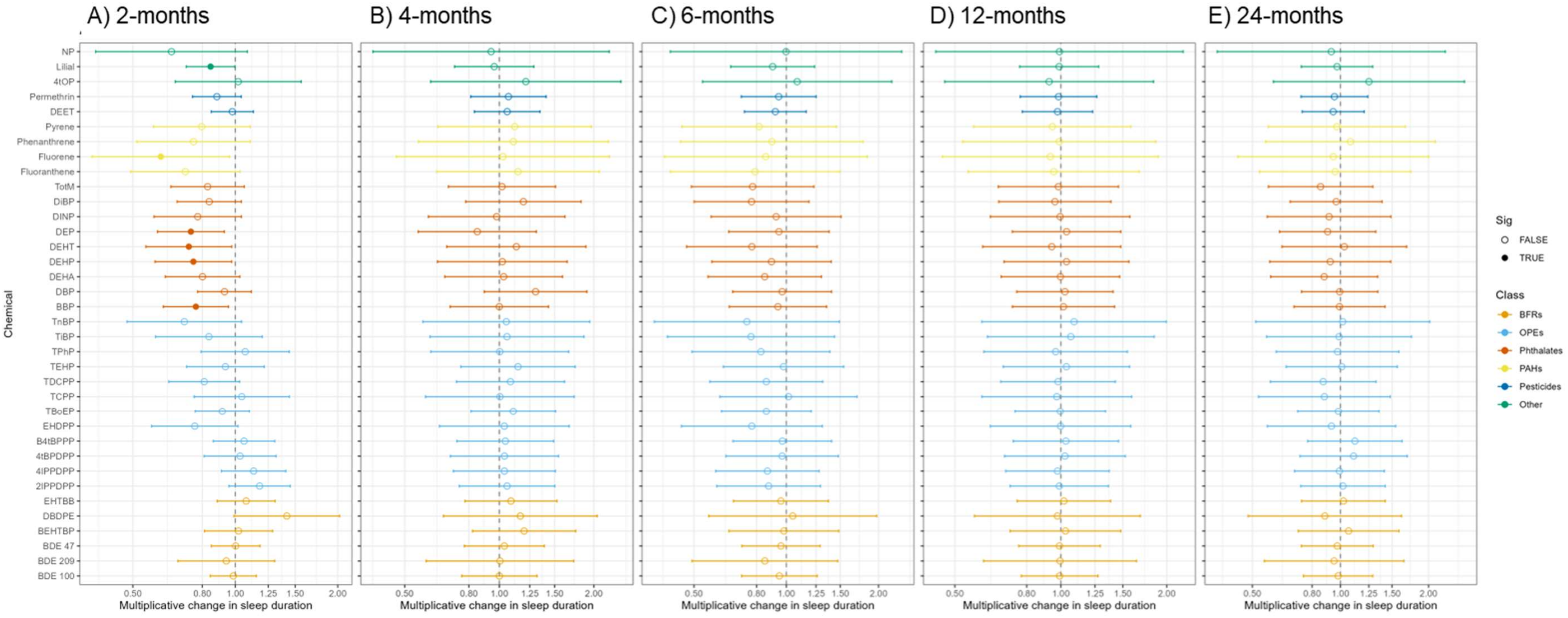
Age-specific estimates from generalized linear mixed effects models with an exposure-age interaction term examining associations between SVOC exposure and nighttime sleep at A) 2-months, B) 4-months, C) 6-months, D) 12-months, and E) 24-months old.

### Total sleep duration

We evaluated associations between SVOC exposures and total sleep duration over a 24-hour period, finding similar associations to nighttime sleep duration. Higher exposure to several SVOCs, including PAHs, phthalates, and OPEs, was associated with shorter total sleep duration at 2-months old, but not at other ages (**Supplemental Figure 1; Supplemental Table 6**). Significant associations ranged from 12% (lilial) to 48% (NP) lower expected sleep per 10-fold increase in SVOC exposure. A 10-fold increase in exposure was significantly associated with lower expected total sleep duration in 2-month-olds across phthalates (BBP, DEP, di-isononyl phthalate (DINP), bis (2-ethylhexyl) terephthalate (DEHT), bis(2-ethylhexyl) phthalate (DEHP); 21–25% lower), OPEs (tri-n-butyl phosphate (TnBP), tris (1,3-dichloro-isopropyl) phosphate (TDCPP), 2-ethylhexyl diphenyl phosphate (EHDPP); 17–31% lower), and PAHs (fluorene, phenanthrene, fluoranthene, and pyrene; 25–44% lower). Total SVOC exposure was associated with a significant decrease in total sleep among 2-month-olds (e^β^=0.79, 95% CI: 0.67, 0.95) while associations at other ages were null (**Supplemental Table 7**).

## Discussion

Prenatal exposure to SVOCs may be associated with adverse effects on sleep duration in early infancy. We observed negative associations between daytime, nighttime, and total sleep duration and prenatal exposures to individual SVOCs and the SVOC mixture among 2-month-old infants. Further, associations between prenatal SVOC exposures and sleep duration were modified by age. These findings suggest prenatal chemical exposures may disrupt initial establishment of sleep patterns in newborns, although effects may attenuate with age.

The strongest associations between SVOC exposures and sleep duration were observed at 2-month assessments, the time point closest to the exposure period. These effects may be most evident during early infancy due to an immature circadian system, evolving sleep architecture, intermittent sleep periods, and large interindividual variability.^2,4,22,23^ Although circadian entrainment begins in utero, external light cues and diurnal hormonal signals conveyed through breast milk during the first two years of life are critical for circadian system maturation.^18^ Changes in sleep needs across early infancy may further be impacted by age-related developmental milestones related to learning new skills and concepts, including object permanence, separation anxiety, and motor skills, which may have a larger impact sleep duration in children over time than prenatal exposures.^4^

While SVOC exposure was associated with significantly decreased expected daytime sleep at 2-months of age, exposure tended to be associated with increased daytime sleep at older ages, albeit non-significant. Although associations differed in direction across age groups, both may indicate adverse effects on age-appropriate sleep duration patterns. Lower expected daytime sleep at 2-months old may indicate insufficient sleep during a period of high sleep need, and increased daytime sleep in older children may suggest persistence of daytime sleep needs during a developmental period where daytime sleep duration is expected to decrease.^2,24^

Several previous studies have identified associations between prenatal phthalate and phenol exposures and sleep problems as measured by parent-report screeners.^13,16,17^ Using data from the National Institutes of Health-funded Environmental influences on Child Health Outcomes (ECHO) Cohort, higher prenatal exposure to selected phenols were generally associated with worse sleep outcomes across parent-report scales of sleep problems among children aged 1.5 to 8 years, though phenol exposure assessed as a mixture was not associated with sleep outcomes.^16,17^ We observed a significant decrease in sleep duration in relation to NP exposure; however, the phenol chemicals included in the ECHO study were not measured in our analysis. The ECHO study also found that child sex was an effect modifier of the association between phenol exposure and sleep outcomes. While we did not stratify by sex in the current study due to the small sample size, future studies will be needed to evaluate this relationship. Phthalate exposures have also been associated with child sleep. Among Taiwanese children aged 1.5 to 3, metabolites of dibutyl phthalate (DBP) and DEP were associated with higher t-scores on a sleep problems survey scale.^13^ We did not identify significant associations between phthalate exposures and sleep duration at 12 and 24 months; however, we observed a significant decrease in sleep duration associated with phthalate exposure at 2- and 4-months of age, including exposure to BBP and DEP.

Our study has several strengths and limitations. Limitations include the use of categorical sleep duration data obtained via parent report, which does not allow for characterization of sleep duration on the scale of minutes. We therefore applied a Poisson model to model the discrete duration values and align with the outcome distribution, rather than assuming normality as needed for linear regression. These models provide estimates of percent changes in expected sleep duration, providing an intuitive interpretation for stakeholders (e.g., parents, clinicians). While various sociodemographic variables may be associated with sleep duration outcomes, we were limited in the ability to adjust for these factors due to the small sample size and risk of overadjustment. Silicone wristband samplers allowed for measurement of many common SVOCs present in the environment, however our analysis is limited in its use of multiple comparisons for individual SVOC exposures and sleep duration values. For the goals of our study, this approach remains useful in characterizing exposure patterns to co-occurring environmental contaminants and generating hypotheses for associations between environmental chemical exposures and childhood sleep. Strengths of the study include the use of sleep duration as an outcome, a specific facet of sleep health that may be more readily understood than sleep problems broadly defined by behavioral questionnaires. Additionally, the structure of our study cohort allows for continued follow up, enabling additional data collection to further investigate childhood sleep outcomes in future research.

## Conclusions

This study is one of the first to examine sleep duration across multiple chemical classes of SVOCs. Prenatal exposure to SVOCs may be associated with a decrease in childhood sleep duration, particularly among 2-month olds, representing an important developmental timepoint for infant sleep. These findings demonstrate the importance of incorporating age at assessment in analyses of environmental exposures and sleep duration, as the effects of prenatal exposures may decrease with increasing age. Future work should collect detailed sleep duration data and other sleep health indicators using validated parent-report questionnaires and objective measures (e.g., actigraphy, polysomnography) to further assess sleep disruptions and problems and how they may persist over time.

## Data Availability

All data produced in the present study are available upon reasonable request to the authors.

## Acknowledgements and Funding

The authors wish to thank the HOPE 1000 participants that made this work possible. We also wish to thank everyone in Duke Hospital’s labor and delivery unit that supported this research. This work was supported by research grants from the National Institute of Environmental Health Sciences (R01ES031419, P42ES010356, R01AI167850, and T32ES021432).

