## Supplemental Information for "Prenatal exposure to semi-volatile organic compounds and early childhood sleep duration"

**Supplemental Table 1. List of all targeted analytes.** See Hoehn et al. (2027) for batch- specific method detection limits.

| Compound Class | Class Abbreviation | Compound Name | Compound Abbr. | CAS Number |
|---|---|---|---|---|
| Brominated Flame Retardants | BFRs | Decabromodiphenyl ethane | DBDPE | 84852-53-9 |
| | | Bis (2-ethyl hexyl)-2,3,4,5-tetrabromophthalate | BEHTBP | 26040-51-7 |
| | | 2-ethyl hexyl-2,3,4,5-tetrabromobenzoate | EHTBB | 183658-27-7 |
| | | Bistribromophenoxyethane | BTBPE | 37853-59-1 |
| | | 2,4,6-tris(2,4,6-tribromophenoxy)-1,3,5-triazine | TTBP-TAZ | 25713-60-4 |
| | | Tetrabromobisphenol A bis(2,3-dibromopropyl ether) | TBBPA-DBPE | 21850-44-2 |
| | | Tris (2,3-dibromopropyl) isocyanurate | TDBPIC | 52434-90-9 |
| | | Octabromotrimethylphenylindane | OBIND | 155613-93-7 |
| | | 2,4,4'-tribromodiphenyl ether | BDE 28 | 41318-75-6 |
| | | 2,2',4,4'-tetrabromodiphenyl ether | BDE 47 | 5436-43-1 |
| | | 2,3',4,4'-tetrabromodiphenyl ether | BDE 66 | 189084-61-5 |
| | | 2,2',3,4,4'-pentabromodiphenyl ether | BDE 85 | 182346-21-0 |
| | | 2,2',4,4',5-pentabromodiphenyl ether | BDE 99 | 60348-60-9 |
| | | 2,2',4,4',6-pentabromodiphenyl ether | BDE 100 | 189084-64-8 |
| | | 2,2',4,4',5,5'-hexabromodiphenyl ether | BDE 153 | 68631-49-2 |
| | | 2,2',4,4',5,6'-hexabromodiphenyl ether | BDE 154 | 207122-15-4 |
| | | 2,2',3,4,4',5',6-heptabromodiphenyl ether | BDE 183 | 207122-16-5 |
| | | Decabromodiphenyl ether | BDE 209 | 1163-19-5 |
| | OPEs | Triethyl phosphate | TEP | 78-40-0 |

| | | | | |
|---|---|---|---|---|
| Organophosphate Esters | | Tri-iso-butyl-phosphate | TiBP | 126-71-6 |
| | | Triisopropyl phosphate | TiPP | 513-02-0 |
| | | Tri-m-cresyl phosphate | TmCP | 563-04-2 |
| | | Tri-n-butyl-phosphate | TnBP | 126-73-8 |
| | | Tri-o-cresyl phosphate | ToCP | 78-30-8 |
| | | Tri-p-cresyl phosphate | TpCP | 78-32-0 |
| | | Tripentyl phosphate | TPeP | 2528-38-3 |
| | | Triphenyl phosphate | TPHP | 115-86-6 |
| | | Tripropyl phosphate | TPrP | 513-08-06 |
| | | 2-Ethylhexyl diphenyl phosphate | EHDPP | 1241-94-7 |
| | | Tris(2-ethylhexyl) phosphate | TEHP | 78-42-2 |
| | | Tri-(2-butoxyethyl)-phosphate | TBOEP | 78-51-3 |
| | | Tris (2-chloro-ethyl) phosphate | TCEP | 115-96-8 |
| | | Tris (1-chloro-isopropyl) phosphate | TCPP | 13674-84-5 |
| | | Tris (2,4-dichloro-isopropyl) phosphate | TDCPP | 13674-87-8 |
| | | Tris(3,5-dimethyl phenyl) phosphate | TDMPP | 9006-37-5 |
| | | 2-Isopropylphenyl diphenyl phosphate | 2IPPDPP | 64532-94-4 |
| | | 3-Isopropylphenyl diphenyl phosphate | 3IPPDPP | 69515-46-4 |
| | | 4-Isopropylphenyl dipheynyl phosphate | 4IPPDPP | 55864-04-5 |
| | | Bis(2-isopropylphenyl) phenyl phosphate | B2IPPPP | 69500-29-4 |
| | | Bis (3-isopropylphenyl) phenyl phosphate | B3IPPPP | 69500-30-7 |
| | | Bis (4-isopopylphenyl) phenyl phosphate | B4IPPPP | 55864-07-8 |
| | | 2,4-Diisopropylphenyl diphenyl phosphate | 24DIPPDPP | n.a. |
| | | Tris(3-isopropylphenyl) phosphate | T3IPPP | 72668-27-0 |
| | | Tris(4-isopropylphenyl) phosphate | T4IPPP | 26967-76-0 |
| | | Bis (2,4-diisopropylphenyl) phenyl phosphate | B24DIPPPP | n.a. |
| | | 2-tert-butylphenyl diphenyl phosphate | 2tBPDPP | 83242-23-3 |
| | | 4-tert-butylphenyl diphenyl phosphate | 4tBPDPP | 981-40-8 |
| | | bis(2-tert-butylphenyl) phenyl phosphate | B2tBPPP | 65652-41-7 |

| | | | | |
|---|---|---|---|---|
| | | bis(4-tert-butylphenyl) phenyl phosphate | B4tBPPP | 115-87-7 |
| Polycyclic Aromatic Hydrocarbons | PAHs | 3-Methylcholanthrene | 3-Methylcholanthrene | 56-49-5 |
| | | 7,12-Dimethylbenz(a)anthracene | 7,12-Dimethylbenz(a)anthracene | 57-97-6 |
| | | Acenaphthylene | Acenaphthylene | 208-96-8 |
| | | Acenapthene | Acenapthene | 83-32-9 |
| | | Anthracene | Anthracene | 120-12-7 |
| | | 1,2 Benzanthracene (Benz[a]anthracene) | Benz[a]anthracene | 56-55-3 |
| | | Benzo(c)phenanthrene | Benzo(c)phenanthrene | 195-19-7 |
| | | Benzo(g,h,i)perylene | Benzo(g,h,i)perylene | 191-24-2 |
| | | Benzo[a]pyrene | Benzo[a]pyrene | 50-32-8 |
| | | Benzo[e]pyrene | Benzo[e]pyrene | 192-97-2 |
| | | Chrysene | Chrysene | 218-01-9 |
| | | Dibenz(a,h)anthracene | Dibenz(a,h)anthracene | 53-70-3 |
| | | Dibenz[a,h]pyrene | Dibenz[a,h]pyrene | 189-64-0 |
| | | Dibenz[a,i]pyrene | Dibenz[a,i]pyrene | 189-55-9 |
| | | Dibenz[a,l]pyrene | Dibenz[a,l]pyrene | 191-30-0 |
| | | Fluoranthene | Fluoranthene | 206-44-0 |
| | | Fluorene | Fluorene | 86-73-7 |
| | | Indeno(1,2,3-cd)pyrene | Indeno(1,2,3-cd)pyrene | 193-39-5 |
| | | Naphthalene | Naphthalene | 91-20-3 |
| | | Perylene | Perylene | 198-55-0 |
| | | Phenanthrene | Phenanthrene | 85-01-8 |
| | | Pyrene | Pyrene | 129-00-0 |

| | | | | |
|---|---|---|---|---|
| Polychlorinated Biphenyls | PCBs | 3,3'-Dichlorobiphenyl | PCB 11 | 2050-67-1 |
| | | 2,4,4'-Trichlorobiphenyl | PCB28 | 7012-37-5 |
| | | 2,2',4,4'-Tetrachlorobiphenyl | PCB47 | 2437-79-8 |
| | | 2,2',4,6'-Tetrachlorobiphenyl | PCB51 | 68491-04-7 |
| | | 2,2',5,5'-Tetrachlorobiphenyl | PCB52 | 35693-99-3 |
| | | 2,3',4,5'-Tetrachlorobiphenyl | PCB68 | 73575-52-7 |
| | | 2,2',4,5,5'-Pentachlorobiphenyl | PCB101 | 37680-73-2 |
| | | 2,3',4,4',5-Pentachlorobiphenyl | PCB118 | 31508-00-6 |
| | | 2,2',3,4,4',5'-Hexachlorobiphenyl | PCB138 | 35065-28-2 |
| | | 2,2',4,4',5,5'-Hexachlorobiphenyl | PCB153 | 35065-27-1 |
| | | 2,2',3,4,4',5,5'-Heptachlorobiphenyl | PCB180 | 35065-29-3 |
| | | 2,2',3,4,4',5',6-Heptachlorobiphenyl | PCB183 | 52663-69-1 |
| Pesticides | Pesticides | Atrazine | Atrazine | 1912-24-9 |
| | | Azoxystrobin | Azoxystrobin | 131860-33-8 |
| | | chlorfenapyr | chlorfenapyr | 122453-73-0 |
| | | Chlorpyrifos | Chlorpyrifos | 2921-88-2 |
| | | cis-Chlordane | cis-Chlordane | 5103-71-9 |
| | | cis-Permethrin | cis-Permethrin | 61949-76-6 |
| | | cypermethrin | cypermethrin | 52315-07-8 |
| | | Fipronil | Fipronil | 120068-37-3 |
| | | Fluoxastrobin | Fluoxastrobin | 193740-76-0 |
| | | DEET | DEET | 134-62-3 |
| | | Lindane | Lindane | 58-89-9 |
| | | Malathion | Malathion | 121-75-5 |
| | | p,p'-DDE | p,p'-DDE | 72-55-9 |
| | | pyraclostrobin | pyraclostrobin | 175013-18-0 |
| | | trans-Chlordane | trans-Chlordane | 5103-74-2 |
| | | trans-Permethrin | trans-Permethrin | 61949-77-7 |

| | | | | |
|---|---|---|---|---|
| | | Trifloxystrobin | Trifloxystrobin | 141517-21-7 |
| | | propiconazole | Pzole | 60207-90-1 |
| | | Cyprodinil | Cyprodinil | 121552-61-2 |
| | | Oxyfluorfen | Oxyfluorfen | 42874-03-3 |
| Phthalates and Phthalate Alternatives | Phthalates | benzyl butyl phthalate | BBP | 85-68-7 |
| | | dibutyl phthalate | DBP | 84-74-2 |
| | | Bis (2-ethylhexyl) adipate | DEHA | 103-23-1 |
| | | Bis(2-ethylhexyl) phthalate | DEHP | 117-81-7 |
| | | Bis (2-ethylhexyl) terephthalate | DEHT | 6422-86-2 |
| | | di-ethyl phthalate | DEP | 84-66-2 |
| | | di-isobutyl phthalate | DiBP | 84-69-5 |
| | | di-isononyl phthalate | DINP | 68515-48-0 |
| | | di-methyl phthalate | DMP | 131-11-3 |
| | | trioctylmetallitate | TOTM | 3319-31-1 |
| Industrial Chemicals | Other | nonylphenol isomer mix | NP | 25154-52-3 |
| | | 4-tertoctylphenol | OP | 27193-28-8 |
| Personal Care Products | Other | lilial | lilial | 80-54-6 |
| | | nicotine | nicotine | 54-11-5 |

**Supplemental Table 2.** Intraclass correlation coefficients (ICCs) for SVOCs detected in at least 70% of wristband samples.

| Class | Chemical | ICC |
|---|---|---|
| **BFRs** | BDE 100 | 0.520 |
| | BDE 209 | 0.633 |
| | BDE 47 | 0.667 |
| | BEHTBP | 0.633 |
| | DBDPE | 0.691 |
| | EHTBB | 0.620 |
| **OPEs** | 2IPPDPP | 0.449 |
| | 4IPPDPP | 0.348 |
| | 4tBPDPP | 0.518 |
| | B4tBPPP | 0.541 |
| | EHDPP | 0.674 |
| | TBoEP | 0.770 |
| | TCPP | 0.602 |
| | TDCPP | 0.642 |
| | TEHP | 0.630 |
| | TiBP | 0.779 |
| | TnBP | 0.750 |
| | TPhP | 0.416 |
| **Other** | 4tOP | 0.869 |
| | Lilial | 0.738 |
| | NP | 0.727 |
| **PAHs** | Fluoranthene | 0.852 |
| | Fluorene | 0.492 |
| | Phenanthrene | 0.656 |
| | Pyrene | 0.707 |
| **Pesticides** | DEET | 0.647 |
| | Permethrin | 0.809 |
| **Phthalates** | BBP | 0.901 |

| | | |
|---|---|---|
| | DBP | 0.651 |
| | DEHA | 0.657 |
| | DEHP | 0.749 |
| | DEHT | 0.745 |
| | DEP | 0.861 |
| | DiBP | 0.723 |
| | DINP | 0.654 |
| | TotM | 0.676 |

**Supplemental Table 3. Detection frequency and summary statistics of all 117 targeted SVOCs in wristbands (n=323).**

| | | Trimester 1 (n=49) | | | Trimester 2 (n=138) | | | Trimester 3 (n=136) | | |
|---|---|---|---|---|---|---|---|---|---|---|
| Class | Chemical | Detection Frequency (%) | Median (ng/g) | 95th Percentile (ng/g) | Detection Frequency (%) | Median (ng/g) | 95th Percentile (ng/g) | Detection Frequency (%) | Median (ng/g) | 95th Percentile (ng/g) |
| BFRs | BDE 100 | 82 | 2.910 | 23.94 | 90 | 3.070 | 32.50 | 86 | 2.378 | 26.06 |
| | BDE 153 | 39 | -- | 12.04 | 51 | 2.353 | 31.55 | 44 | -- | 14.13 |
| | BDE 154 | 59 | 2.119 | 19.83 | 56 | 2.169 | 25.44 | 48 | -- | 13.49 |
| | BDE 183 | 8 | -- | 2.583 | 17 | -- | 1.527 | 15 | -- | 2.130 |
| | BDE 209 | 100 | 4.564 | 14.67 | 99 | 4.587 | 22.04 | 98 | 4.517 | 22.69 |
| | BDE 28 | 49 | -- | 4.123 | 49 | -- | 4.889 | 44 | -- | 3.332 |
| | BDE 47 | 94 | 16.63 | 126.6 | 97 | 20.15 | 274.1 | 96 | 15.28 | 155.6 |
| | BDE 66 | 22 | -- | 4.519 | 42 | -- | 8.174 | 32 | -- | 6.913 |
| | BDE 85 | 31 | -- | 11.40 | 46 | -- | 19.69 | 35 | -- | 12.06 |
| | BDE 99 | 61 | 32.36 | 180.5 | 66 | 37.09 | 417.8 | 60 | 22.83 | 300.9 |
| | BEHTBP | 100 | 30.47 | 352.2 | 100 | 30.93 | 357.7 | 99 | 37.97 | 333.6 |
| | BTBPE | 2 | -- | -- | 7 | -- | 14.16 | 5 | -- | 13.78 |
| | DBDPE | 59 | 13.44 | 34.06 | 75 | 9.140 | 32.87 | 72 | 9.457 | 35.55 |
| | EHTBB | 90 | 53.32 | 397.5 | 93 | 55.63 | 885.0 | 93 | 65.12 | 815.0 |
| | OBIND | 0 | -- | -- | 1 | -- | -- | 0 | -- | -- |
| | TDBPIC | 2 | -- | -- | 5 | -- | 2.116 | 2 | -- | -- |
| | TTBP-TAZ | 0 | -- | -- | 4 | -- | -- | 3 | -- | -- |
| OPEs | 24DIPPDPP | 37 | -- | 210.5 | 63 | 38.10 | 193.5 | 56 | 31.33 | 310.4 |
| | 2IPPDPP | 100 | 39.09 | 377.6 | 95 | 87.37 | 492.9 | 94 | 43.72 | 810.4 |
| | 2tBPDPP | 0 | -- | -- | 1 | -- | -- | 0 | -- | -- |
| | 3IPPDPP | 39 | -- | 65.90 | 56 | 25.96 | 157.6 | 45 | -- | 126.0 |
| | 4IPPDPP | 96 | 26.90 | 231.3 | 90 | 63.38 | 432.5 | 88 | 39.18 | 466.2 |
| | 4tBPDPP | 98 | 28.44 | 229.1 | 100 | 48.26 | 396.7 | 99 | 46.26 | 297.9 |

| | | | | | | | | | | |
|---|---|---|---|---|---|---|---|---|---|---|
| | B24DIPPPP | 0 | -- | -- | 0 | -- | -- | 0 | -- | -- |
| | B2IPPPP | 57 | 32.97 | 170.3 | 70 | 66.31 | 297.6 | 57 | 49.69 | 326.9 |
| | B2tBPPP | 0 | -- | -- | 0 | -- | -- | 0 | -- | -- |
| | B3IPPPP | 2 | -- | -- | 8 | -- | 21.98 | 3 | -- | -- |
| | B4IPPPP | 59 | 5.030 | 50.42 | 72 | 15.72 | 107.3 | 64 | 11.09 | 108.3 |
| | B4tBPPP | 94 | 14.20 | 89.76 | 91 | 23.72 | 182.4 | 90 | 26.15 | 153.4 |
| | EHDPP | 100 | 34.79 | 252.35 | 100 | 42.15 | 243.0 | 100 | 38.96 | 234.0 |
| | T3IPPP | 0 | -- | -- | 0 | -- | -- | 0 | -- | -- |
| | T4IPPP | 0 | -- | -- | 1 | -- | -- | 1 | -- | -- |
| | TBoEP | 96 | 167.5 | 759.4 | 96 | 169.3 | 1047 | 93 | 148.2 | 1508 |
| | TCEP | 53 | 7.438 | 63.22 | 36 | -- | 172.7 | 36 | -- | 264.9 |
| | TCPP | 100 | 157.1 | 663.0 | 100 | 175.8 | 863.6 | 100 | 154.8 | 749.1 |
| | TDCPP | 100 | 205.8 | 1964 | 100 | 187.0 | 1726 | 99 | 204.5 | 1066 |
| | TDMPP | 12 | -- | 8.427 | 30 | -- | 33.14 | 20 | -- | 18.95 |
| | TEHP | 100 | 26.52 | 185.6 | 99 | 21.05 | 128.9 | 99 | 21.15 | 103.6 |
| | TEP | 69 | 20.70 | 549.5 | 51 | 28.00 | 215.8 | 60 | 24.62 | 275.2 |
| | TiBP | 92 | 4.364 | 21.62 | 93 | 4.483 | 20.80 | 96 | 4.036 | 25.01 |
| | TiPP | 0 | -- | -- | 0 | -- | -- | 0 | -- | -- |
| | TmCP | 8 | -- | 7.996 | 3 | -- | -- | 4 | -- | -- |
| | TnBP | 84 | 21.94 | 64.50 | 94 | 19.20 | 61.52 | 93 | 18.66 | 49.08 |
| | ToCP | 0 | -- | -- | 0 | -- | -- | 0 | -- | -- |
| | TpCP | 0 | -- | -- | 1 | -- | -- | 1 | -- | -- |
| | TPeP | 0 | -- | -- | 0 | -- | -- | 0 | -- | -- |
| | TPhP | 100 | 119.9 | 574.0 | 100 | 240.6 | 924.1 | 100 | 201.3 | 1411 |
| | TPrP | 0 | -- | -- | 0 | -- | -- | 0 | -- | -- |
| Other | 4tOP | 100 | 20.84 | 50.32 | 100 | 15.91 | 55.32 | 100 | 15.65 | 53.37 |
| | Lilial | 100 | 633.0 | 5879 | 100 | 673.5 | 8014 | 100 | 488.2 | 7991 |
| | Nicotine | 12 | -- | 378.9 | 9 | -- | 7213 | 7 | -- | 11228 |
| | NP | 100 | 364.0 | 801.7 | 100 | 266.3 | 696.9 | 100 | 260.5 | 825.8 |

| | | | | | | | | | | |
|---|---|---|---|---|---|---|---|---|---|---|
| PAHs | 3-Methylcholanthrene | 0 | -- | -- | 0 | -- | -- | 0 | -- | -- |
| | 7,12-Dimethyl benz(a)anthracene | 0 | -- | -- | 0 | -- | -- | 0 | -- | -- |
| | Acenaphthylene | 37 | -- | 8.459 | 28 | -- | 6.379 | 28 | -- | 6.366 |
| | Acenapthene | 51 | 10.65 | 98.17 | 42 | -- | 29.38 | 43 | -- | 39.70 |
| | Anthracene | 14 | -- | 39.49 | 17 | -- | 16.09 | 13 | -- | 36.10 |
| | Benz[a]anthracene | 16 | -- | 13.94 | 17 | -- | 7.064 | 10 | -- | 14.18 |
| | Benzo(c)phenanthrene | 2 | -- | -- | 4 | -- | -- | 4 | -- | -- |
| | Benzo(g,h,i)perylene | 0 | -- | -- | 3 | -- | -- | 3 | -- | -- |
| | Benzo[a]pyrene | 4 | -- | -- | 4 | -- | -- | 4 | -- | -- |
| | Benzo[e]pyrene | 8 | -- | 3.504 | 14 | -- | 3.140 | 10 | -- | 6.024 |
| | Benzo[j,b,k]fluoranthene | 12 | -- | 12.67 | 19 | -- | 8.967 | 13 | -- | 16.55 |
| | Chrysene | 35 | -- | 8.312 | 33 | -- | 9.376 | 25 | -- | 20.52 |
| | Dibenz(a,h)anthracene | 0 | -- | -- | 1 | -- | -- | 1 | -- | -- |
| | Dibenz[a,h]pyrene | 0 | -- | -- | 0 | -- | -- | 0 | -- | -- |
| | Dibenz[a,i]pyrene | 0 | -- | -- | 0 | -- | -- | 0 | -- | -- |
| | Dibenz[a,l]pyrene | 0 | -- | -- | 0 | -- | -- | 0 | -- | -- |
| | Fluoranthene | 98 | 8.341 | 27.58 | 97 | 8.584 | 29.75 | 98 | 7.394 | 30.79 |
| | Fluorene | 84 | 3.803 | 11.27 | 72 | 3.495 | 18.99 | 76 | 3.369 | 14.22 |

| | | | | | | | | | | |
|---|---|---|---|---|---|---|---|---|---|---|
| | Indeno(1,2,3-cd)pyrene | 4 | -- | -- | 4 | -- | -- | 1 | -- | -- |
| | Perylene | 2 | -- | -- | 2 | -- | -- | 2 | -- | -- |
| | Phenanthrene | 92 | 26.76 | 59.09 | 96 | 27.15 | 76.06 | 91 | 26.20 | 86.87 |
| | Naphthalene | 6 | -- | 164.9 | 12 | -- | 905.6 | 13 | -- | 5197 |
| | Pyrene | 100 | 6.826 | 24.37 | 93 | 6.800 | 25.72 | 94 | 6.416 | 22.81 |
| | PCB 101 | 0 | -- | -- | 7 | -- | 14.19 | 7 | -- | 24.33 |
| | PCB 11 | 37 | -- | 2.162 | 61 | 0.6380 | 1.606 | 60 | 0.6375 | 1.448 |
| | PCB 118 | 0 | -- | -- | 4 | -- | -- | 4 | -- | -- |
| | PCB 138 | 0 | -- | -- | 1 | -- | -- | 3 | -- | -- |
| | PCB 153 | 0 | -- | -- | 0 | -- | -- | 2 | -- | -- |
| PCBs | PCB 180 | 0 | -- | -- | 1 | -- | -- | 1 | -- | -- |
| | PCB 183 | 0 | -- | -- | 0 | -- | -- | 0 | -- | -- |
| | PCB 28 | 10 | -- | 12.40 | 13 | -- | 24.97 | 13 | -- | 31.22 |
| | PCB 47 | 4 | -- | -- | 4 | -- | -- | 3 | -- | -- |
| | PCB 51 | 0 | -- | -- | 0 | -- | -- | 0 | -- | -- |
| | PCB 52 | 2 | -- | -- | 7 | -- | 31.78 | 7 | -- | 34.44 |
| | PCB 68 | 2 | -- | -- | 3 | -- | -- | 3 | -- | -- |
| | Atrazine | 0 | -- | -- | 0 | -- | -- | 0 | -- | -- |
| | Azoxystrobin | 33 | -- | 163.0 | 25 | -- | 97.20 | 15 | -- | 95.61 |
| | Chlorfenapyr | 0 | -- | -- | 3 | -- | -- | 2 | -- | -- |
| | Chlorpyrifos | 6 | -- | 6.303 | 5 | -- | 670.7 | 2 | -- | -- |
| | cis-Chlordane | 24 | -- | 51.71 | 33 | -- | 48.97 | 35 | -- | 49.88 |
| | cis-Permethrin | 98 | 22.05 | 591.6 | 99 | 28.99 | 752.7 | 99 | 26.81 | 469.8 |
| Pesticides | Cypermethrin | 16 | -- | 168.5 | 22 | -- | 656.4 | 20 | -- | 900.0 |
| | Cyprodinil | 0 | -- | -- | 3 | -- | -- | 1 | -- | -- |
| | DEET | 98 | 288.0 | 18003 | 99 | 246.6 | 4291 | 99 | 284.7 | 2235 |
| | Fipronil | 14 | -- | 142.9 | 15 | -- | 989.0 | 10 | -- | 371.7 |
| | Fluoxastrobin | 0 | -- | -- | 0 | -- | -- | 0 | -- | -- |
| | Lindane | 6 | -- | 8.298 | 3 | -- | -- | 4 | -- | -- |
| | Malathion | 2 | -- | -- | 0 | -- | -- | 0 | -- | -- |

| | | | | | | | | | | |
|---|---|---|---|---|---|---|---|---|---|---|
| | Oxyfluorfen | 0 | -- | -- | 0 | -- | -- | 0 | -- | -- |
| | p,p'-DDE | 6 | -- | 3.373 | 7 | -- | 8.923 | 10 | -- | 8.850 |
| | Propiconazole | 0 | -- | -- | 0 | -- | -- | 1 | -- | -- |
| | Pyraclostrobin | 0 | -- | -- | 1 | -- | -- | 0 | -- | -- |
| | trans-Chlordane | 55 | 7.239 | 78.46 | 49 | -- | 86.16 | 51 | 7.560 | 109.7 |
| | trans-Permethrin | 98 | 32.12 | 724.1 | 99 | 38.24 | 672.7 | 99 | 29.65 | 515.2 |
| | Trifloxystrobin | 0 | -- | -- | 0 | -- | -- | 0 | -- | -- |
| Phthalates | BBP | 100 | 168.6 | 2348 | 100 | 231.4 | 2547 | 99 | 230.3 | 2254 |
| | DBP | 100 | 1109 | 3877 | 100 | 1129 | 4965 | 100 | 1153 | 7176 |
| | DEHA | 100 | 2976 | 59686 | 100 | 5269 | 45393 | 100 | 4583 | 36174 |
| | DEHP | 100 | 1289 | 5427 | 100 | 1488 | 6738 | 100 | 1295 | 9870 |
| | DEHT | 100 | 83577 | 1468896 | 100 | 110215 | 820821 | 100 | 101352 | 703815 |
| | DEP | 100 | 412.7 | 2850 | 100 | 517.4 | 8973 | 100 | 447.0 | 3152 |
| | DiBP | 65 | 1018 | 4593 | 86 | 1220 | 6817 | 80 | 1155 | 4825 |
| | DINP | 100 | 57305 | 425700 | 100 | 53297 | 347142 | 100 | 48803 | 328765 |
| | DMP | 24 | -- | 82.93 | 20 | -- | 238.0 | 21 | -- | 173.8 |
| | TotM | 100 | 1722 | 13581 | 100 | 2173 | 12707 | 100 | 2161 | 12130 |

**Supplemental Table 4. Prenatal exposure to SVOCs detected in at least 70% of wristbands.** Exposures were averaged across all trimester wristbands per participant.

| Class | Chemical | Minimum (ng/g) | 25th Percentile (ng/g) | Median (ng/g) | 75th Percentile (ng/g) | 95th Percentile (ng/g) | Maximum (ng/g) |
|---|---|---|---|---|---|---|---|
| **BFRs** | BDE 100 | 0.0985 | 1.055 | 2.566 | 7.454 | 27.27 | 101.8 |
| | BDE 209 | 0.6542 | 2.970 | 4.950 | 8.466 | 25.99 | 240.7 |
| | BDE 47 | 0.4671 | 8.114 | 20.68 | 47.15 | 173.8 | 570.8 |
| | BEHTBP | 2.406 | 16.58 | 39.61 | 87.26 | 345.9 | 497.0 |
| | DBDPE | 1.041 | 4.274 | 6.516 | 13.06 | 30.79 | 63.21 |
| | EHTBB | 3.366 | 26.10 | 60.52 | 133.5 | 705.2 | 1876 |
| **OPEs** | 2IPPDPP | 3.166 | 27.84 | 66.20 | 127.8 | 574.3 | 1574 |
| | 4IPPDPP | 1.871 | 19.66 | 50.21 | 106.6 | 372.3 | 556.2 |
| | 4tBPDPP | 1.360 | 27.38 | 61.59 | 107.2 | 376.4 | 1212 |
| | B4tBPPP | 0.7436 | 9.797 | 24.43 | 49.96 | 172.7 | 699.0 |
| | EHDPP | 3.429 | 27.25 | 45.14 | 69.99 | 278.3 | 761.9 |
| | TBoEP | 4.251 | 75.60 | 174.0 | 327.6 | 974.1 | 12220 |
| | TCPP | 21.09 | 109.8 | 175.1 | 294.1 | 736.0 | 7452 |
| | TDCPP | 22.47 | 110.3 | 220.7 | 494.4 | 1389 | 5249 |
| | TEHP | 0.7614 | 13.52 | 22.11 | 48.42 | 129.8 | 510.0 |
| | TiBP | 0.4570 | 2.491 | 3.839 | 7.920 | 21.64 | 51.66 |
| | TnBP | 2.501 | 12.13 | 18.31 | 30.36 | 51.74 | 265.7 |
| | TPhP | 26.01 | 133.3 | 226.9 | 379.5 | 1106 | 3993 |
| **Other** | 4tOP | 2.569 | 11.27 | 15.93 | 26.52 | 49.72 | 91.01 |
| | Lilial | 12.72 | 222.5 | 662.7 | 1868 | 8215 | 19184 |
| | NP | 90.98 | 192.9 | 265.0 | 377.0 | 612.4 | 1451 |
| **PAHs** | Fluoranthene | 0.6412 | 5.195 | 8.140 | 14.10 | 28.85 | 207.5 |
| | Fluorene | 0.4418 | 2.527 | 4.334 | 5.058 | 10.36 | 26.23 |
| | Phenanthrene | 4.737 | 18.00 | 26.94 | 37.07 | 81.71 | 169.0 |
| | Pyrene | 0.7328 | 4.432 | 6.717 | 11.10 | 22.96 | 133.4 |
| **Pesticides** | DEET | 11.65 | 108.7 | 252.6 | 909.6 | 10801 | 396440 |
| | Permethrin | 2.057 | 29.36 | 58.86 | 238.5 | 993.5 | 19104 |
| **Phthalates** | BBP | 30.19 | 121.7 | 226.4 | 677.5 | 2358 | 23803 |
| | DBP | 118.6 | 711.8 | 1186 | 2134 | 6732 | 2693752 |
| | DEHA | 849.1 | 2816 | 5170 | 10549 | 43741 | 136753 |
| | DEHP | 153.5 | 741.3 | 1515 | 2689 | 8537 | 37456 |
| | DEHT | 15077 | 71184 | 100427 | 232184 | 791399 | 2075109 |
| | DEP | 42.85 | 290.0 | 535.9 | 1102 | 5309 | 100688 |
| | DiBP | 123.6 | 552.5 | 995.2 | 1884 | 5144 | 485708 |
| | DINP | 8514.9 | 30158 | 55958 | 110686 | 310826 | 1068837 |
| | TotM | 164.7 | 1056 | 2379 | 4474 | 13144 | 93503 |

**Supplemental Table 5. Spearman correlations coefficients ($r_s$) for average prenatal exposures among mother-child pairs (n = 147)**

| | | | | | | | | | | | | | | | | | | | | | | | | | | | | | | | | | | | | |
|---|---|---|---|---|---|---|---|---|---|---|---|---|---|---|---|---|---|---|---|---|---|---|---|---|---|---|---|---|---|---|---|---|---|---|---|---|
| TiBP | 1.00 | | | | | | | | | | | | | | | | | | | | | | | | | | | | | | | | | | | |
| TnBP | 0.31 | 1.00 | | | | | | | | | | | | | | | | | | | | | | | | | | | | | | | | | | |
| TCPP | 0.19 | 0.26 | 1.00 | | | | | | | | | | | | | | | | | | | | | | | | | | | | | | | | | |
| TDCPP | 0.12 | 0.25 | 0.28 | 1.00 | | | | | | | | | | | | | | | | | | | | | | | | | | | | | | | | |
| TBoEP | 0.29 | 0.38 | 0.14 | 0.27 | 1.00 | | | | | | | | | | | | | | | | | | | | | | | | | | | | | | | |
| TPhP | 0.09 | 0.14 | 0.15 | 0.34 | 0.22 | 1.00 | | | | | | | | | | | | | | | | | | | | | | | | | | | | | | |
| EHDPP | 0.22 | 0.38 | 0.38 | 0.40 | 0.41 | 0.35 | 1.00 | | | | | | | | | | | | | | | | | | | | | | | | | | | | | |
| TEHP | 0.34 | 0.28 | 0.10 | 0.25 | 0.30 | 0.11 | 0.32 | 1.00 | | | | | | | | | | | | | | | | | | | | | | | | | | | | |
| 2IPPDPP | 0.05 | -0.03 | 0.07 | 0.15 | 0.09 | 0.72 | 0.18 | 0.13 | 1.00 | | | | | | | | | | | | | | | | | | | | | | | | | | | |
| 4IPPDPP | 0.05 | 0.01 | 0.10 | 0.25 | 0.12 | 0.75 | 0.19 | 0.17 | 0.92 | 1.00 | | | | | | | | | | | | | | | | | | | | | | | | | | |
| 4tBPDPP | 0.01 | 0.20 | 0.21 | 0.46 | 0.30 | 0.39 | 0.39 | 0.31 | 0.18 | 0.29 | 1.00 | | | | | | | | | | | | | | | | | | | | | | | | | |
| B4tBPPP | 0.01 | 0.17 | 0.20 | 0.42 | 0.31 | 0.39 | 0.35 | 0.31 | 0.22 | 0.32 | 0.96 | 1.00 | | | | | | | | | | | | | | | | | | | | | | | | |
| Fluorene | 0.24 | 0.05 | 0.06 | 0.20 | 0.13 | 0.14 | 0.23 | 0.12 | 0.01 | 0.01 | 0.10 | 0.07 | 1.00 | | | | | | | | | | | | | | | | | | | | | | | |
| Phenanthrene | 0.25 | 0.29 | 0.17 | 0.15 | 0.17 | 0.00 | 0.33 | 0.15 | -0.05 | -0.08 | 0.18 | 0.17 | 0.61 | 1.00 | | | | | | | | | | | | | | | | | | | | | | |
| Fluoranthene | 0.22 | 0.28 | 0.19 | 0.36 | 0.40 | 0.18 | 0.45 | 0.38 | -0.01 | 0.09 | 0.40 | 0.38 | 0.44 | 0.62 | 1.00 | | | | | | | | | | | | | | | | | | | | | |
| Pyrene | 0.20 | 0.30 | 0.24 | 0.47 | 0.43 | 0.24 | 0.49 | 0.35 | 0.07 | 0.14 | 0.43 | 0.42 | 0.47 | 0.56 | 0.85 | 1.00 | | | | | | | | | | | | | | | | | | | | |
| DEET | 0.13 | 0.11 | 0.23 | 0.19 | 0.15 | 0.20 | 0.34 | 0.18 | 0.06 | 0.16 | 0.28 | 0.24 | 0.27 | 0.15 | 0.30 | 0.34 | 1.00 | | | | | | | | | | | | | | | | | | | |
| DEP | 0.24 | 0.19 | 0.15 | 0.16 | 0.16 | 0.11 | 0.35 | 0.10 | -0.12 | -0.09 | 0.21 | 0.20 | 0.28 | 0.35 | 0.27 | 0.35 | 0.21 | 1.00 | | | | | | | | | | | | | | | | | | |
| DBP | 0.19 | 0.14 | 0.15 | 0.09 | 0.08 | 0.07 | 0.35 | 0.18 | 0.03 | 0.03 | 0.07 | 0.10 | 0.05 | 0.21 | 0.16 | 0.22 | 0.10 | 0.18 | 1.00 | | | | | | | | | | | | | | | | | |
| DiBP | 0.27 | 0.34 | 0.34 | 0.38 | 0.36 | 0.29 | 0.54 | 0.22 | 0.07 | 0.16 | 0.41 | 0.41 | 0.23 | 0.50 | 0.56 | 0.54 | 0.23 | 0.49 | 0.43 | 1.00 | | | | | | | | | | | | | | | | |
| BBP | 0.13 | 0.44 | 0.25 | 0.24 | 0.39 | 0.17 | 0.43 | 0.16 | 0.09 | 0.13 | 0.23 | 0.24 | 0.15 | 0.28 | 0.26 | 0.34 | 0.23 | 0.25 | 0.20 | 0.48 | 1.00 | | | | | | | | | | | | | | | |
| DEHA | 0.25 | 0.42 | 0.29 | 0.43 | 0.45 | 0.31 | 0.46 | 0.32 | 0.12 | 0.16 | 0.34 | 0.31 | 0.21 | 0.16 | 0.31 | 0.40 | 0.19 | 0.32 | 0.15 | 0.41 | 0.36 | 1.00 | | | | | | | | | | | | | | |
| DEHP | 0.29 | 0.37 | 0.30 | 0.34 | 0.32 | 0.24 | 0.52 | 0.27 | 0.08 | 0.16 | 0.42 | 0.40 | 0.27 | 0.34 | 0.38 | 0.44 | 0.37 | 0.34 | 0.30 | 0.55 | 0.51 | 0.46 | 1.00 | | | | | | | | | | | | | |
| DEHT | 0.18 | 0.40 | 0.29 | 0.38 | 0.43 | 0.29 | 0.56 | 0.39 | 0.12 | 0.15 | 0.39 | 0.39 | 0.19 | 0.19 | 0.41 | 0.49 | 0.16 | 0.28 | 0.23 | 0.49 | 0.35 | 0.62 | 0.41 | 1.00 | | | | | | | | | | | | |
| DINP | 0.09 | 0.20 | 0.27 | 0.49 | 0.20 | 0.34 | 0.49 | 0.35 | 0.15 | 0.22 | 0.48 | 0.45 | 0.24 | 0.15 | 0.31 | 0.36 | 0.22 | 0.22 | 0.09 | 0.32 | 0.22 | 0.33 | 0.40 | 0.46 | 1.00 | | | | | | | | | | | |
| TotM | 0.13 | 0.22 | 0.34 | 0.27 | 0.21 | 0.34 | 0.34 | 0.32 | 0.13 | 0.19 | 0.26 | 0.21 | 0.12 | 0.19 | 0.29 | 0.28 | 0.26 | 0.21 | 0.12 | 0.39 | 0.20 | 0.33 | 0.35 | 0.41 | 0.47 | 1.00 | | | | | | | | | | |
| 4tOP | 0.23 | -0.02 | 0.07 | -0.03 | -0.08 | -0.05 | 0.13 | 0.17 | 0.18 | 0.10 | -0.10 | -0.11 | 0.18 | 0.21 | 0.15 | 0.13 | 0.03 | -0.12 | 0.17 | 0.09 | -0.04 | -0.06 | 0.05 | 0.00 | 0.12 | -0.08 | 1.00 | | | | | | | | | |
| NP | 0.30 | 0.11 | 0.22 | 0.26 | 0.13 | 0.17 | 0.39 | 0.31 | 0.17 | 0.14 | 0.09 | 0.04 | 0.32 | 0.29 | 0.22 | 0.26 | 0.03 | 0.09 | 0.27 | 0.16 | 0.23 | 0.22 | 0.33 | 0.26 | 0.41 | 0.25 | 0.34 | 1.00 | | | | | | | | |
| Lilial | 0.13 | 0.15 | 0.01 | 0.15 | 0.04 | 0.04 | 0.02 | 0.09 | -0.15 | -0.02 | 0.23 | 0.20 | 0.22 | 0.16 | 0.21 | 0.19 | 0.01 | 0.30 | 0.00 | 0.12 | 0.12 | 0.13 | 0.17 | 0.20 | 0.23 | 0.19 | -0.23 | 0.17 | 1.00 | | | | | | | |
| BDE 47 | 0.07 | 0.12 | 0.18 | 0.22 | 0.22 | 0.24 | 0.41 | 0.16 | 0.22 | 0.32 | 0.26 | 0.28 | 0.10 | 0.21 | 0.31 | 0.29 | 0.22 | 0.09 | 0.20 | 0.33 | 0.30 | 0.02 | 0.38 | 0.18 | 0.31 | 0.19 | 0.11 | 0.26 | 0.17 | 1.00 | | | | | | |
| BDE 100 | 0.09 | 0.13 | 0.17 | 0.25 | 0.24 | 0.25 | 0.44 | 0.19 | 0.20 | 0.31 | 0.27 | 0.29 | 0.10 | 0.19 | 0.32 | 0.32 | 0.18 | 0.14 | 0.21 | 0.34 | 0.31 | 0.04 | 0.39 | 0.23 | 0.35 | 0.22 | 0.11 | 0.31 | 0.18 | 0.96 | 1.00 | | | | | |
| EHTBB | -0.01 | 0.03 | 0.20 | 0.20 | 0.23 | 0.20 | 0.21 | 0.12 | 0.18 | 0.28 | 0.42 | 0.47 | 0.02 | 0.04 | 0.24 | 0.22 | 0.28 | 0.04 | -0.05 | 0.19 | 0.30 | 0.23 | 0.28 | 0.18 | 0.19 | 0.10 | -0.03 | -0.03 | 0.07 | 0.28 | 0.26 | 1.00 | | | | |
| BEHTBP | 0.18 | 0.14 | 0.15 | 0.22 | 0.31 | 0.23 | 0.29 | 0.32 | 0.15 | 0.25 | 0.53 | 0.53 | 0.05 | 0.10 | 0.33 | 0.36 | 0.38 | 0.11 | 0.10 | 0.32 | 0.31 | 0.30 | 0.35 | 0.29 | 0.30 | 0.21 | 0.08 | 0.09 | 0.12 | 0.22 | 0.21 | 0.68 | 1.00 | | | |
| BDE 209 | -0.04 | 0.12 | 0.09 | 0.34 | 0.24 | 0.13 | 0.20 | 0.12 | 0.13 | 0.20 | 0.22 | 0.26 | 0.11 | 0.13 | 0.31 | 0.35 | 0.16 | 0.02 | 0.06 | 0.32 | 0.29 | 0.13 | 0.28 | 0.31 | 0.19 | 0.21 | -0.08 | -0.03 | 0.04 | 0.29 | 0.28 | 0.23 | 0.18 | 1.00 | | |
| DBDPE | -0.01 | 0.07 | 0.00 | 0.03 | 0.07 | 0.08 | -0.01 | 0.01 | 0.11 | 0.13 | 0.08 | 0.10 | -0.01 | -0.07 | 0.04 | 0.06 | 0.17 | -0.18 | -0.01 | -0.04 | -0.09 | -0.18 | 0.00 | -0.06 | 0.02 | -0.07 | -0.02 | -0.18 | -0.15 | -0.02 | -0.01 | 0.17 | 0.10 | 0.11 | 1.00 | |
| Permethrin | 0.11 | 0.14 | 0.10 | 0.16 | 0.23 | 0.17 | 0.32 | 0.35 | 0.06 | 0.12 | 0.14 | 0.13 | 0.02 | 0.02 | 0.28 | 0.27 | 0.35 | 0.07 | 0.16 | 0.20 | 0.09 | 0.12 | 0.28 | 0.16 | 0.21 | 0.25 | 0.11 | 0.14 | -0.05 | 0.30 | 0.28 | 0.14 | 0.20 | 0.20 | 0.11 | 1.00 |
| | TiBP | TnBP | TCPP | TDCPP | TBoEP | TPhP | EHDPP | TEHP | 2IPPDPP | 4IPPDPP | 4tBPDPP | B4tBPPP | Fluorene | Phenanthrene | Fluoranthene | Pyrene | DEET | DEP | DBP | DiBP | BBP | DEHA | DEHP | DEHT | DINP | TotM | 4tOP | NP | Lilial | BDE 47 | BDE 100 | EHTBB | BEHTBP | BDE 209 | DBDPE | Permethrin |

**Supplemental Table 6. Results from generalized linear mixed effects models with an exposure-age interaction term examining associations between SVOC exposure and daytime, nighttime, and total sleep duration.** Results are the multiplicative change in sleep duration (hours) per 10-fold increase in SVOC exposure.

| | | | **Daytime** | | **Nighttime** | | **Total** | |
|---|---|---|---|---|---|---|---|---|
| **Class** | **Chemical** | **Age** | **$e^{\beta}$ (95% CI)** | **p-value** | **$e^{\beta}$ (95% CI)** | **p-value** | **$e^{\beta}$ (95% CI)** | **p-value** |
| BFRs | BDE 100 | 2 | 0.98 (0.83, 1.17) | 0.83 | 0.99 (0.84, 1.15) | 0.86 | 0.99 (0.88, 1.11) | 0.83 |
| | | 4 | 1.20 (0.86, 1.69) | 0.28 | 1.00 (0.76, 1.32) | 1.00 | 1.07 (0.86, 1.32) | 0.54 |
| | | 6 | 1.02 (0.70, 1.49) | 0.92 | 0.95 (0.71, 1.26) | 0.72 | 0.97 (0.78, 1.21) | 0.78 |
| | | 12 | 1.39 (0.98, 1.96) | 0.06 | 0.99 (0.77, 1.27) | 0.95 | 1.06 (0.87, 1.28) | 0.57 |
| | | 24 | 0.99 (0.62, 1.57) | 0.96 | 0.98 (0.75, 1.29) | 0.89 | 0.98 (0.79, 1.22) | 0.88 |
| | BDE 209 | 2 | 0.91 (0.63, 1.31) | 0.60 | 0.94 (0.68, 1.31) | 0.71 | 0.93 (0.73, 1.18) | 0.53 |
| | | 4 | 0.87 (0.45, 1.68) | 0.68 | 1.00 (0.58, 1.73) | 0.99 | 0.96 (0.64, 1.45) | 0.84 |
| | | 6 | 0.97 (0.49, 1.94) | 0.93 | 0.85 (0.49, 1.47) | 0.56 | 0.89 (0.58, 1.35) | 0.57 |
| | | 12 | 1.58 (0.85, 2.94) | 0.15 | 0.99 (0.60, 1.64) | 0.98 | 1.10 (0.75, 1.61) | 0.63 |
| | | 24 | 1.00 (0.41, 2.43) | 0.99 | 0.95 (0.55, 1.65) | 0.86 | 0.96 (0.62, 1.48) | 0.85 |
| | BDE 47 | 2 | 0.99 (0.82, 1.19) | 0.91 | 1.00 (0.85, 1.18) | 0.98 | 1.00 (0.89, 1.13) | 0.99 |
| | | 4 | 1.24 (0.86, 1.79) | 0.25 | 1.04 (0.77, 1.39) | 0.81 | 1.11 (0.88, 1.38) | 0.39 |
| | | 6 | 1.03 (0.70, 1.52) | 0.88 | 0.96 (0.72, 1.29) | 0.79 | 0.98 (0.78, 1.23) | 0.86 |
| | | 12 | 1.38 (0.95, 2.01) | 0.09 | 0.99 (0.76, 1.29) | 0.95 | 1.06 (0.86, 1.30) | 0.60 |
| | | 24 | 1.03 (0.65, 1.65) | 0.90 | 0.98 (0.74, 1.29) | 0.86 | 0.98 (0.78, 1.23) | 0.89 |
| | BEHTBP | 2 | 1.01 (0.77, 1.31) | 0.96 | 1.02 (0.81, 1.29) | 0.85 | 1.02 (0.86, 1.21) | 0.80 |
| | | 4 | 1.16 (0.73, 1.83) | 0.53 | 1.20 (0.82, 1.75) | 0.35 | 1.19 (0.89, 1.58) | 0.24 |
| | | 6 | 0.96 (0.56, 1.66) | 0.88 | 0.98 (0.65, 1.48) | 0.93 | 0.98 (0.71, 1.35) | 0.88 |
| | | 12 | 1.12 (0.69, 1.82) | 0.66 | 1.03 (0.72, 1.48) | 0.87 | 1.04 (0.79, 1.38) | 0.76 |
| | | 24 | 0.85 (0.43, 1.66) | 0.63 | 1.07 (0.72, 1.58) | 0.75 | 1.03 (0.75, 1.42) | 0.86 |
| | DBDPE | 2 | 1.16 (0.77, 1.75) | 0.48 | 1.42 (0.99, 2.03) | 0.06 | 1.28 (0.98, 1.67) | 0.07 |
| | | 4 | 1.07 (0.54, 2.10) | 0.86 | 1.17 (0.66, 2.05) | 0.59 | 1.13 (0.74, 1.73) | 0.57 |
| | | 6 | 1.35 (0.59, 3.11) | 0.48 | 1.05 (0.56, 1.97) | 0.88 | 1.12 (0.69, 1.83) | 0.64 |
| | | 12 | 1.28 (0.64, 2.58) | 0.48 | 0.98 (0.57, 1.68) | 0.94 | 1.04 (0.69, 1.57) | 0.86 |
| | | 24 | 1.21 (0.46, 3.19) | 0.70 | 0.88 (0.48, 1.62) | 0.69 | 0.94 (0.58, 1.51) | 0.79 |

| | | | | | | | | |
|---|---|---|---|---|---|---|---|---|
| | EHTBB | 2 | 0.98 (0.78, 1.22) | 0.84 | 1.08 (0.88, 1.31) | 0.46 | 1.03 (0.89, 1.19) | 0.69 |
| | | 4 | 0.92 (0.60, 1.41) | 0.70 | 1.09 (0.78, 1.53) | 0.62 | 1.03 (0.80, 1.34) | 0.82 |
| | | 6 | 1.00 (0.62, 1.60) | 0.98 | 0.96 (0.67, 1.37) | 0.82 | 0.97 (0.73, 1.28) | 0.83 |
| | | 12 | 1.22 (0.81, 1.84) | 0.34 | 1.02 (0.75, 1.38) | 0.90 | 1.06 (0.83, 1.34) | 0.65 |
| | | 24 | 0.78 (0.46, 1.33) | 0.36 | 1.02 (0.74, 1.42) | 0.89 | 0.99 (0.76, 1.28) | 0.92 |
| OPEs | 2IPPDPP | 2 | 0.95 (0.75, 1.19) | 0.66 | 1.18 (0.96, 1.45) | 0.12 | 1.06 (0.91, 1.24) | 0.45 |
| | | 4 | 1.24 (0.81, 1.91) | 0.32 | 1.06 (0.75, 1.50) | 0.75 | 1.12 (0.86, 1.46) | 0.41 |
| | | 6 | 0.86 (0.51, 1.46) | 0.58 | 0.88 (0.59, 1.29) | 0.50 | 0.87 (0.64, 1.18) | 0.38 |
| | | 12 | 1.04 (0.68, 1.58) | 0.86 | 0.99 (0.72, 1.37) | 0.95 | 1.00 (0.78, 1.28) | 1.00 |
| | | 24 | 0.82 (0.49, 1.36) | 0.45 | 1.02 (0.73, 1.42) | 0.91 | 0.99 (0.76, 1.28) | 0.93 |
| | 4IPPDPP | 2 | 0.98 (0.77, 1.24) | 0.84 | 1.13 (0.91, 1.41) | 0.26 | 1.06 (0.90, 1.24) | 0.50 |
| | | 4 | 1.30 (0.82, 2.05) | 0.27 | 1.04 (0.71, 1.51) | 0.85 | 1.12 (0.84, 1.49) | 0.44 |
| | | 6 | 0.85 (0.51, 1.41) | 0.53 | 0.87 (0.59, 1.28) | 0.47 | 0.86 (0.64, 1.17) | 0.34 |
| | | 12 | 1.24 (0.80, 1.92) | 0.34 | 0.98 (0.70, 1.37) | 0.90 | 1.03 (0.79, 1.33) | 0.85 |
| | | 24 | 0.80 (0.46, 1.39) | 0.43 | 0.99 (0.70, 1.41) | 0.96 | 0.96 (0.73, 1.27) | 0.78 |
| | 4tBPDPP | 2 | 0.88 (0.67, 1.16) | 0.36 | 1.03 (0.81, 1.32) | 0.80 | 0.97 (0.81, 1.16) | 0.71 |
| | | 4 | 1.15 (0.71, 1.85) | 0.57 | 1.04 (0.70, 1.54) | 0.85 | 1.08 (0.80, 1.45) | 0.63 |
| | | 6 | 1.08 (0.63, 1.87) | 0.77 | 0.97 (0.64, 1.48) | 0.88 | 1.00 (0.72, 1.39) | 1.00 |
| | | 12 | 1.50 (0.87, 2.59) | 0.14 | 1.03 (0.69, 1.52) | 0.89 | 1.10 (0.81, 1.50) | 0.52 |
| | | 24 | 1.11 (0.55, 2.23) | 0.78 | 1.11 (0.73, 1.69) | 0.63 | 1.09 (0.78, 1.54) | 0.60 |
| | B4tBPPP | 2 | 0.91 (0.72, 1.15) | 0.43 | 1.06 (0.86, 1.31) | 0.58 | 0.99 (0.85, 1.15) | 0.91 |
| | | 4 | 1.05 (0.68, 1.61) | 0.82 | 1.05 (0.73, 1.49) | 0.81 | 1.05 (0.80, 1.37) | 0.73 |
| | | 6 | 1.09 (0.67, 1.76) | 0.73 | 0.97 (0.67, 1.41) | 0.87 | 1.00 (0.75, 1.33) | 0.99 |
| | | 12 | 1.54 (0.96, 2.50) | 0.08 | 1.03 (0.73, 1.46) | 0.85 | 1.12 (0.85, 1.46) | 0.42 |
| | | 24 | 1.04 (0.55, 1.96) | 0.92 | 1.12 (0.77, 1.62) | 0.55 | 1.10 (0.81, 1.48) | 0.55 |
| | EHDPP | 2 | 0.65 (0.48, 0.90) | 0.01 | 0.76 (0.57, 1.02) | 0.07 | 0.71 (0.58, 0.89) | 0.00 |
| | | 4 | 1.09 (0.62, 1.89) | 0.77 | 1.04 (0.64, 1.67) | 0.88 | 1.06 (0.74, 1.51) | 0.77 |
| | | 6 | 1.27 (0.65, 2.47) | 0.49 | 0.77 (0.45, 1.31) | 0.34 | 0.90 (0.60, 1.35) | 0.60 |
| | | 12 | 1.08 (0.60, 1.97) | 0.79 | 1.00 (0.63, 1.58) | 0.99 | 1.01 (0.71, 1.44) | 0.93 |
| | | 24 | 1.20 (0.53, 2.73) | 0.66 | 0.93 (0.56, 1.54) | 0.78 | 0.96 (0.64, 1.44) | 0.86 |
| | TBoEP | 2 | 0.94 (0.77, 1.16) | 0.58 | 0.92 (0.76, 1.10) | 0.35 | 0.93 (0.82, 1.07) | 0.31 |

| | | | | | | | | |
|---|---|---|---|---|---|---|---|---|
| | | 4 | 1.25 (0.87, 1.81) | 0.23 | 1.11 (0.81, 1.51) | 0.52 | 1.16 (0.92, 1.46) | 0.22 |
| | | 6 | 1.32 (0.85, 2.06) | 0.21 | 0.86 (0.61, 1.21) | 0.38 | 0.98 (0.75, 1.27) | 0.86 |
| | | 12 | 1.00 (0.67, 1.48) | 0.99 | 1.00 (0.74, 1.34) | 0.98 | 1.00 (0.79, 1.25) | 0.98 |
| | | 24 | 1.00 (0.59, 1.70) | 1.00 | 0.98 (0.71, 1.35) | 0.92 | 0.98 (0.76, 1.27) | 0.88 |
| | TCPP | 2 | 0.87 (0.60, 1.27) | 0.48 | 1.04 (0.76, 1.44) | 0.79 | 0.97 (0.76, 1.23) | 0.80 |
| | | 4 | 1.07 (0.54, 2.12) | 0.84 | 1.00 (0.58, 1.73) | 0.99 | 1.03 (0.68, 1.56) | 0.90 |
| | | 6 | 1.13 (0.59, 2.17) | 0.71 | 1.02 (0.61, 1.70) | 0.95 | 1.05 (0.71, 1.56) | 0.82 |
| | | 12 | 1.23 (0.66, 2.32) | 0.52 | 0.97 (0.60, 1.59) | 0.92 | 1.02 (0.70, 1.49) | 0.91 |
| | | 24 | 1.19 (0.53, 2.66) | 0.67 | 0.88 (0.52, 1.48) | 0.63 | 0.92 (0.61, 1.39) | 0.69 |
| | TDCPP | 2 | 0.83 (0.64, 1.09) | 0.19 | 0.81 (0.64, 1.03) | 0.08 | 0.83 (0.69, 0.99) | 0.04 |
| | | 4 | 1.07 (0.67, 1.73) | 0.77 | 1.09 (0.73, 1.61) | 0.69 | 1.08 (0.80, 1.46) | 0.60 |
| | | 6 | 1.01 (0.58, 1.77) | 0.96 | 0.86 (0.56, 1.31) | 0.49 | 0.90 (0.65, 1.25) | 0.54 |
| | | 12 | 1.29 (0.79, 2.11) | 0.30 | 0.98 (0.68, 1.43) | 0.92 | 1.04 (0.78, 1.38) | 0.80 |
| | | 24 | 1.25 (0.62, 2.50) | 0.53 | 0.87 (0.58, 1.32) | 0.52 | 0.92 (0.66, 1.28) | 0.61 |
| | TEHP | 2 | 0.87 (0.65, 1.16) | 0.34 | 0.94 (0.72, 1.22) | 0.62 | 0.91 (0.75, 1.10) | 0.32 |
| | | 4 | 1.02 (0.62, 1.68) | 0.95 | 1.15 (0.75, 1.74) | 0.52 | 1.10 (0.80, 1.51) | 0.55 |
| | | 6 | 1.17 (0.65, 2.11) | 0.61 | 0.98 (0.62, 1.54) | 0.93 | 1.03 (0.73, 1.46) | 0.87 |
| | | 12 | 1.52 (0.87, 2.66) | 0.14 | 1.04 (0.69, 1.57) | 0.86 | 1.12 (0.81, 1.53) | 0.50 |
| | | 24 | 1.34 (0.66, 2.73) | 0.42 | 1.01 (0.65, 1.56) | 0.96 | 1.05 (0.74, 1.48) | 0.80 |
| | TiBP | 2 | 1.03 (0.69, 1.54) | 0.89 | 0.84 (0.58, 1.20) | 0.33 | 0.93 (0.71, 1.22) | 0.60 |
| | | 4 | 1.09 (0.56, 2.12) | 0.79 | 1.06 (0.60, 1.86) | 0.85 | 1.07 (0.70, 1.63) | 0.76 |
| | | 6 | 1.62 (0.75, 3.51) | 0.22 | 0.77 (0.41, 1.44) | 0.41 | 0.97 (0.60, 1.57) | 0.90 |
| | | 12 | 0.75 (0.37, 1.48) | 0.40 | 1.07 (0.62, 1.84) | 0.81 | 0.99 (0.66, 1.50) | 0.98 |
| | | 24 | 1.00 (0.43, 2.32) | 0.99 | 0.99 (0.56, 1.75) | 0.97 | 0.99 (0.63, 1.54) | 0.96 |
| | TnBP | 2 | 0.63 (0.40, 0.98) | 0.04 | 0.71 (0.48, 1.04) | 0.08 | 0.69 (0.52, 0.92) | 0.01 |
| | | 4 | 1.33 (0.65, 2.74) | 0.43 | 1.05 (0.57, 1.94) | 0.87 | 1.15 (0.73, 1.82) | 0.55 |
| | | 6 | 1.15 (0.45, 2.92) | 0.78 | 0.74 (0.37, 1.49) | 0.40 | 0.84 (0.49, 1.45) | 0.53 |
| | | 12 | 1.19 (0.53, 2.64) | 0.67 | 1.09 (0.60, 1.99) | 0.78 | 1.10 (0.69, 1.76) | 0.67 |
| | | 24 | 1.02 (0.31, 3.40) | 0.97 | 1.02 (0.51, 2.02) | 0.96 | 1.01 (0.58, 1.77) | 0.97 |
| | TPhP | 2 | 1.01 (0.72, 1.42) | 0.95 | 1.07 (0.79, 1.44) | 0.66 | 1.04 (0.83, 1.29) | 0.74 |
| | | 4 | 1.25 (0.67, 2.30) | 0.48 | 1.00 (0.61, 1.66) | 0.99 | 1.08 (0.74, 1.59) | 0.69 |

| | | | | | | | | |
|---|---|---|---|---|---|---|---|---|
| | | 6 | 0.95 (0.49, 1.85) | 0.87 | 0.83 (0.49, 1.39) | 0.47 | 0.86 (0.58, 1.29) | 0.47 |
| | | 12 | 1.08 (0.58, 2.02) | 0.80 | 0.97 (0.60, 1.55) | 0.89 | 0.99 (0.69, 1.42) | 0.95 |
| | | 24 | 0.82 (0.38, 1.76) | 0.61 | 0.98 (0.60, 1.58) | 0.93 | 0.95 (0.65, 1.40) | 0.81 |
| Other | 4tOP | 2 | 0.84 (0.52, 1.36) | 0.48 | 1.02 (0.67, 1.56) | 0.93 | 0.93 (0.68, 1.28) | 0.66 |
| | | 4 | 1.49 (0.64, 3.47) | 0.36 | 1.21 (0.60, 2.44) | 0.59 | 1.30 (0.76, 2.21) | 0.33 |
| | | 6 | 1.15 (0.46, 2.88) | 0.76 | 1.08 (0.53, 2.21) | 0.82 | 1.10 (0.64, 1.91) | 0.72 |
| | | 12 | 0.72 (0.28, 1.86) | 0.50 | 0.93 (0.47, 1.83) | 0.83 | 0.88 (0.52, 1.50) | 0.65 |
| | | 24 | 1.35 (0.37, 4.95) | 0.65 | 1.25 (0.59, 2.65) | 0.56 | 1.26 (0.68, 2.31) | 0.47 |
| | Lilial | 2 | 0.89 (0.73, 1.08) | 0.23 | 0.85 (0.72, 1.00) | 0.05 | 0.88 (0.78, 0.99) | 0.04 |
| | | 4 | 0.85 (0.59, 1.22) | 0.37 | 0.96 (0.72, 1.29) | 0.80 | 0.92 (0.74, 1.15) | 0.47 |
| | | 6 | 1.17 (0.76, 1.80) | 0.49 | 0.90 (0.66, 1.24) | 0.52 | 0.97 (0.76, 1.25) | 0.82 |
| | | 12 | 1.10 (0.78, 1.56) | 0.58 | 0.99 (0.76, 1.28) | 0.93 | 1.01 (0.83, 1.23) | 0.92 |
| | | 24 | 0.81 (0.51, 1.29) | 0.37 | 0.97 (0.73, 1.29) | 0.85 | 0.94 (0.75, 1.18) | 0.60 |
| | NP | 2 | 0.38 (0.21, 0.68) | 0.00 | 0.65 (0.39, 1.08) | 0.10 | 0.52 (0.36, 0.77) | 0.00 |
| | | 4 | 1.75 (0.63, 4.87) | 0.29 | 0.94 (0.40, 2.24) | 0.89 | 1.18 (0.61, 2.29) | 0.61 |
| | | 6 | 1.75 (0.58, 5.24) | 0.32 | 1.00 (0.42, 2.38) | 1.00 | 1.18 (0.60, 2.31) | 0.63 |
| | | 12 | 0.78 (0.26, 2.32) | 0.65 | 0.99 (0.44, 2.23) | 0.98 | 0.94 (0.50, 1.76) | 0.85 |
| | | 24 | 1.71 (0.37, 7.96) | 0.49 | 0.93 (0.38, 2.28) | 0.87 | 1.01 (0.49, 2.10) | 0.98 |
| PAHs | Fluoranthene | 2 | 0.69 (0.46, 1.04) | 0.07 | 0.71 (0.49, 1.03) | 0.07 | 0.71 (0.54, 0.94) | 0.01 |
| | | 4 | 1.23 (0.61, 2.50) | 0.56 | 1.15 (0.63, 2.08) | 0.65 | 1.18 (0.75, 1.84) | 0.48 |
| | | 6 | 1.28 (0.56, 2.92) | 0.56 | 0.79 (0.42, 1.50) | 0.47 | 0.91 (0.55, 1.49) | 0.70 |
| | | 12 | 1.25 (0.62, 2.51) | 0.53 | 0.95 (0.55, 1.67) | 0.87 | 1.01 (0.66, 1.54) | 0.96 |
| | | 24 | 1.78 (0.71, 4.45) | 0.22 | 0.96 (0.53, 1.74) | 0.89 | 1.04 (0.65, 1.66) | 0.87 |
| | Fluorene | 2 | 0.51 (0.31, 0.84) | 0.01 | 0.60 (0.38, 0.96) | 0.03 | 0.56 (0.40, 0.78) | 0.00 |
| | | 4 | 1.15 (0.46, 2.87) | 0.77 | 1.03 (0.47, 2.24) | 0.95 | 1.07 (0.59, 1.93) | 0.83 |
| | | 6 | 1.46 (0.59, 3.62) | 0.42 | 0.86 (0.40, 1.84) | 0.69 | 1.01 (0.57, 1.80) | 0.98 |
| | | 12 | 0.64 (0.27, 1.52) | 0.31 | 0.93 (0.46, 1.89) | 0.85 | 0.86 (0.51, 1.47) | 0.59 |
| | | 24 | 0.84 (0.27, 2.56) | 0.75 | 0.95 (0.45, 2.00) | 0.88 | 0.92 (0.51, 1.65) | 0.77 |
| | Phenanthrene | 2 | 0.49 (0.32, 0.74) | 0.00 | 0.75 (0.51, 1.11) | 0.15 | 0.62 (0.47, 0.83) | 0.00 |
| | | 4 | 1.27 (0.55, 2.94) | 0.58 | 1.11 (0.55, 2.22) | 0.77 | 1.16 (0.68, 1.99) | 0.58 |
| | | 6 | 1.21 (0.49, 2.94) | 0.68 | 0.90 (0.45, 1.78) | 0.76 | 0.98 (0.57, 1.67) | 0.93 |

| | | | | | | | | |
|---|---|---|---|---|---|---|---|---|
| | | 12 | 0.68 (0.28, 1.64) | 0.39 | 0.99 (0.53, 1.86) | 0.97 | 0.92 (0.56, 1.50) | 0.73 |
| | | 24 | 2.04 (0.66, 6.35) | 0.22 | 1.08 (0.56, 2.11) | 0.82 | 1.18 (0.69, 2.02) | 0.55 |
| | Pyrene | 2 | 0.68 (0.48, 0.98) | 0.04 | 0.80 (0.57, 1.11) | 0.17 | 0.75 (0.59, 0.95) | 0.02 |
| | | 4 | 1.18 (0.60, 2.32) | 0.64 | 1.12 (0.64, 1.96) | 0.70 | 1.14 (0.74, 1.75) | 0.55 |
| | | 6 | 1.17 (0.55, 2.49) | 0.68 | 0.82 (0.46, 1.46) | 0.49 | 0.90 (0.58, 1.42) | 0.66 |
| | | 12 | 1.26 (0.65, 2.44) | 0.50 | 0.95 (0.57, 1.58) | 0.83 | 1.00 (0.68, 1.48) | 0.99 |
| | | 24 | 1.14 (0.49, 2.67) | 0.75 | 0.97 (0.57, 1.66) | 0.92 | 0.98 (0.64, 1.50) | 0.93 |
| Pesticides | DEET | 2 | 1.04 (0.89, 1.22) | 0.64 | 0.98 (0.85, 1.13) | 0.78 | 1.00 (0.90, 1.11) | 0.99 |
| | | 4 | 1.04 (0.78, 1.40) | 0.78 | 1.06 (0.83, 1.35) | 0.64 | 1.05 (0.88, 1.26) | 0.58 |
| | | 6 | 0.98 (0.74, 1.31) | 0.92 | 0.92 (0.73, 1.16) | 0.49 | 0.94 (0.79, 1.12) | 0.49 |
| | | 12 | 1.18 (0.87, 1.60) | 0.29 | 0.98 (0.78, 1.23) | 0.85 | 1.02 (0.85, 1.22) | 0.83 |
| | | 24 | 1.39 (0.97, 1.99) | 0.07 | 0.94 (0.74, 1.21) | 0.65 | 1.01 (0.83, 1.23) | 0.91 |
| | Permethrin | 2 | 1.06 (0.89, 1.27) | 0.50 | 0.88 (0.75, 1.04) | 0.14 | 0.96 (0.85, 1.08) | 0.50 |
| | | 4 | 1.21 (0.87, 1.66) | 0.25 | 1.07 (0.81, 1.41) | 0.64 | 1.11 (0.90, 1.37) | 0.31 |
| | | 6 | 0.96 (0.68, 1.37) | 0.83 | 0.94 (0.71, 1.25) | 0.69 | 0.95 (0.77, 1.18) | 0.64 |
| | | 12 | 1.15 (0.85, 1.56) | 0.38 | 0.98 (0.77, 1.27) | 0.91 | 1.02 (0.84, 1.23) | 0.86 |
| | | 24 | 1.24 (0.86, 1.79) | 0.26 | 0.95 (0.73, 1.24) | 0.72 | 1.00 (0.81, 1.22) | 0.97 |
| Phthalates | BBP | 2 | 0.73 (0.57, 0.93) | 0.01 | 0.77 (0.61, 0.95) | 0.02 | 0.75 (0.64, 0.89) | 0.00 |
| | | 4 | 1.19 (0.78, 1.81) | 0.42 | 1.00 (0.70, 1.43) | 1.00 | 1.07 (0.81, 1.40) | 0.65 |
| | | 6 | 1.03 (0.66, 1.62) | 0.89 | 0.94 (0.65, 1.35) | 0.74 | 0.97 (0.73, 1.28) | 0.81 |
| | | 12 | 1.20 (0.79, 1.80) | 0.39 | 1.02 (0.73, 1.42) | 0.92 | 1.05 (0.82, 1.36) | 0.69 |
| | | 24 | 1.28 (0.76, 2.17) | 0.35 | 0.99 (0.69, 1.42) | 0.97 | 1.03 (0.78, 1.36) | 0.84 |
| | DBP | 2 | 1.01 (0.84, 1.23) | 0.89 | 0.93 (0.78, 1.11) | 0.43 | 0.96 (0.85, 1.10) | 0.59 |
| | | 4 | 1.17 (0.73, 1.89) | 0.52 | 1.30 (0.90, 1.90) | 0.17 | 1.26 (0.95, 1.68) | 0.11 |
| | | 6 | 1.16 (0.71, 1.89) | 0.56 | 0.97 (0.67, 1.40) | 0.87 | 1.02 (0.77, 1.36) | 0.88 |
| | | 12 | 1.16 (0.73, 1.82) | 0.53 | 1.03 (0.75, 1.41) | 0.87 | 1.05 (0.82, 1.34) | 0.69 |
| | | 24 | 1.00 (0.62, 1.59) | 0.99 | 0.99 (0.74, 1.34) | 0.97 | 0.99 (0.79, 1.25) | 0.95 |
| | DEHA | 2 | 0.86 (0.65, 1.15) | 0.32 | 0.80 (0.62, 1.03) | 0.08 | 0.84 (0.70, 1.01) | 0.07 |
| | | 4 | 0.69 (0.40, 1.19) | 0.18 | 1.03 (0.67, 1.59) | 0.89 | 0.91 (0.65, 1.26) | 0.57 |
| | | 6 | 1.03 (0.60, 1.76) | 0.92 | 0.85 (0.56, 1.30) | 0.46 | 0.90 (0.65, 1.25) | 0.53 |
| | | 12 | 1.19 (0.72, 1.95) | 0.50 | 1.00 (0.68, 1.47) | 0.99 | 1.03 (0.77, 1.38) | 0.84 |

| | | | | | | | | |
|---|---|---|---|---|---|---|---|---|
| | | 24 | 1.23 (0.64, 2.36) | 0.54 | 0.88 (0.58, 1.34) | 0.55 | 0.92 (0.66, 1.28) | 0.61 |
| | DEHP | 2 | 0.79 (0.59, 1.05) | 0.10 | 0.75 (0.58, 0.98) | 0.03 | 0.78 (0.64, 0.94) | 0.01 |
| | | 4 | 1.33 (0.74, 2.37) | 0.34 | 1.02 (0.64, 1.64) | 0.93 | 1.12 (0.78, 1.61) | 0.54 |
| | | 6 | 1.03 (0.59, 1.81) | 0.92 | 0.89 (0.57, 1.40) | 0.63 | 0.93 (0.66, 1.32) | 0.69 |
| | | 12 | 1.29 (0.75, 2.19) | 0.36 | 1.04 (0.69, 1.56) | 0.86 | 1.08 (0.79, 1.48) | 0.63 |
| | | 24 | 1.74 (0.77, 3.93) | 0.19 | 0.92 (0.57, 1.49) | 0.74 | 1.01 (0.68, 1.48) | 0.97 |
| | DEHT | 2 | 0.85 (0.63, 1.16) | 0.31 | 0.73 (0.55, 0.98) | 0.03 | 0.79 (0.64, 0.97) | 0.03 |
| | | 4 | 0.90 (0.49, 1.66) | 0.74 | 1.13 (0.68, 1.89) | 0.63 | 1.05 (0.71, 1.54) | 0.80 |
| | | 6 | 1.15 (0.64, 2.08) | 0.64 | 0.77 (0.47, 1.26) | 0.30 | 0.87 (0.60, 1.26) | 0.47 |
| | | 12 | 1.41 (0.79, 2.49) | 0.24 | 0.94 (0.60, 1.48) | 0.80 | 1.02 (0.73, 1.43) | 0.91 |
| | | 24 | 1.12 (0.51, 2.45) | 0.77 | 1.03 (0.63, 1.68) | 0.90 | 1.04 (0.70, 1.52) | 0.86 |
| | DEP | 2 | 0.84 (0.66, 1.06) | 0.15 | 0.74 (0.59, 0.93) | 0.01 | 0.79 (0.67, 0.93) | 0.00 |
| | | 4 | 0.57 (0.33, 0.97) | 0.04 | 0.85 (0.55, 1.31) | 0.46 | 0.74 (0.53, 1.04) | 0.08 |
| | | 6 | 1.22 (0.79, 1.87) | 0.37 | 0.95 (0.65, 1.38) | 0.77 | 1.03 (0.78, 1.36) | 0.85 |
| | | 12 | 1.01 (0.64, 1.58) | 0.97 | 1.04 (0.73, 1.48) | 0.83 | 1.03 (0.79, 1.35) | 0.84 |
| | | 24 | 1.33 (0.77, 2.32) | 0.31 | 0.90 (0.62, 1.32) | 0.60 | 0.95 (0.71, 1.28) | 0.76 |
| | DiBP | 2 | 0.99 (0.79, 1.23) | 0.91 | 0.84 (0.67, 1.04) | 0.11 | 0.91 (0.78, 1.06) | 0.21 |
| | | 4 | 1.37 (0.83, 2.28) | 0.22 | 1.19 (0.78, 1.82) | 0.41 | 1.26 (0.91, 1.73) | 0.16 |
| | | 6 | 1.16 (0.68, 1.97) | 0.59 | 0.77 (0.50, 1.19) | 0.24 | 0.88 (0.63, 1.22) | 0.44 |
| | | 12 | 1.18 (0.72, 1.93) | 0.52 | 0.96 (0.67, 1.39) | 0.84 | 1.00 (0.76, 1.33) | 0.98 |
| | | 24 | 1.37 (0.85, 2.20) | 0.20 | 0.97 (0.67, 1.39) | 0.85 | 1.03 (0.78, 1.35) | 0.85 |
| | DINP | 2 | 0.71 (0.51, 0.99) | 0.04 | 0.78 (0.58, 1.04) | 0.09 | 0.75 (0.60, 0.93) | 0.01 |
| | | 4 | 1.08 (0.59, 1.99) | 0.79 | 0.98 (0.59, 1.62) | 0.94 | 1.02 (0.70, 1.49) | 0.93 |
| | | 6 | 1.17 (0.64, 2.14) | 0.61 | 0.93 (0.57, 1.51) | 0.76 | 0.99 (0.68, 1.44) | 0.97 |
| | | 12 | 1.41 (0.79, 2.53) | 0.25 | 1.00 (0.63, 1.57) | 0.98 | 1.06 (0.75, 1.51) | 0.72 |
| | | 24 | 1.23 (0.58, 2.64) | 0.59 | 0.91 (0.56, 1.49) | 0.72 | 0.95 (0.65, 1.39) | 0.78 |
| | TotM | 2 | 0.91 (0.69, 1.20) | 0.51 | 0.83 (0.65, 1.06) | 0.14 | 0.87 (0.72, 1.04) | 0.13 |
| | | 4 | 1.03 (0.65, 1.64) | 0.90 | 1.02 (0.69, 1.51) | 0.92 | 1.02 (0.76, 1.37) | 0.88 |
| | | 6 | 0.95 (0.51, 1.76) | 0.86 | 0.78 (0.49, 1.23) | 0.28 | 0.82 (0.57, 1.18) | 0.28 |
| | | 12 | 1.63 (0.95, 2.77) | 0.07 | 0.98 (0.66, 1.46) | 0.94 | 1.08 (0.80, 1.47) | 0.60 |
| | | 24 | 1.32 (0.71, 2.46) | 0.38 | 0.86 (0.57, 1.29) | 0.46 | 0.91 (0.66, 1.26) | 0.57 |

**Supplemental Table 7. Results from generalized linear mixed effects models examining associations between SVOC exposures as an average z-score value and sleep duration outcomes.** Results are the multiplicative change in sleep duration (hours) for a 1 standard deviation increase in z-scored SVOC exposure.

| | **Sleep Duration Outcome** | **Daytime Sleep** | **Nighttime Sleep** | **Total Sleep** |
|---|---|---|---|---|
| **Overall** | **$e^{\beta}$ (95% CI)** | 1.08 (0.93, 1.25) | 0.95 (0.86, 1.04) | 0.99 (0.91, 1.07) |
| | **p-value** | 0.32 | 0.28 | 0.72 |
| **2-months** | **$e^{\beta}$ (95% CI)** | 0.77 (0.59, 1.00) | 0.81 (0.64, 1.02) | 0.79 (0.67, 0.95) |
| | **p-value** | 0.05 | 0.08 | 0.01 |
| **4-month** | **$e^{\beta}$ (95% CI)** | 1.22 (0.76, 1.97) | 1.12 (0.75, 1.67) | 1.16 (0.86, 1.57) |
| | **p-value** | 0.40 | 0.57 | 0.34 |
| **6-months** | **$e^{\beta}$ (95% CI)** | 1.14 (0.70, 1.85) | 0.86 (0.58, 1.28) | 0.94 (0.69, 1.27) |
| | **p-value** | 0.59 | 0.46 | 0.67 |
| **12-months** | **$e^{\beta}$ (95% CI)** | 1.40 (0.86, 2.27) | 0.99 (0.68, 1.44) | 1.06 (0.80, 1.41) |
| | **p-value** | 0.17 | 0.96 | 0.67 |
| **24-months** | **$e^{\beta}$ (95% CI)** | 1.27 (0.66, 2.44) | 0.94 (0.63, 1.41) | 0.98 (0.71, 1.35) |
| | **p-value** | 0.47 | 0.78 | 0.90 |
| | **global p-value** | 0.03 | 0.26 | 0.03 |

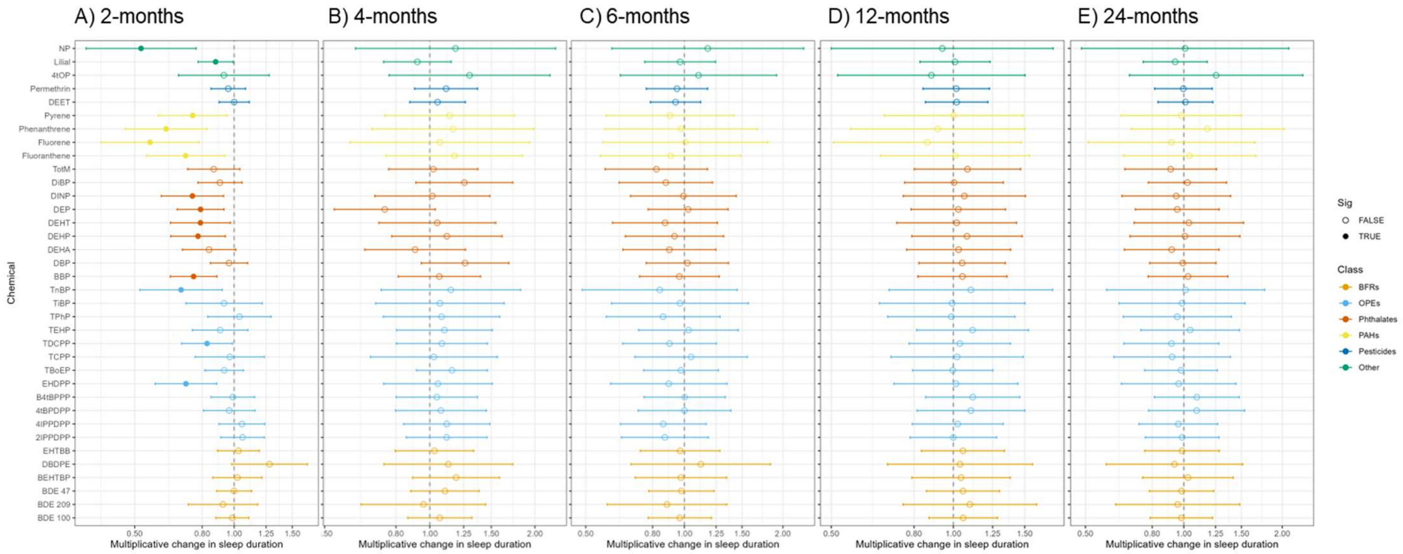


**Supplemental Figure 1. Poisson model for total sleep across a 24-hour period with an exposure-age interaction term. Results presented for age-specific estimates of the effect of SVOCs on sleep duration at A) 2-months, B) 4-months, C) 6-months, D) 12-months, and E) 24-months old.**
